# Early Bedside Biomolecular Phenotyping and Precision Medicine in Lung Transplant Recipients

**DOI:** 10.1101/2025.11.26.25340722

**Authors:** Vittorio Scaravilli, Fabiana Madotto, Sebastiano Maria Colombo, Gloria Turconi, Valentina Vago, Davide Carrà, Marco Bosone, Margherita Brivio, Virginia Beltrama, Letizia Corinna Morlacchi, Leonardo Terranova, Margherita Carnevale-Schianca, Elena Trombetta, Lorenzo Rosso, Alberto Zanella, Mario Nosotti, Francesco Blasi, Lieuwe DJ Bos, Giacomo Grasselli

**Author notes:** **Corresponding author** Dr. Sebastiano Maria Colombo Department of Anesthesia, Critical Care and Emergency, Fondazione IRCCS Ca’ Granda - Ospedale Maggiore Policlinico, Milan, Italy Via F. Sforza, 35, 20122, Milan, Italy Department of Biomedical, Surgical, and Dental Sciences, University of Milan, Milan, Italy Università di Milano, Milan, Italy Via Della Commenda 9, 20122, Milan, Italy.

## Abstract

**Background:** Lung transplantation (LUTX) is frequently complicated by Primary Graft Dysfunction (PGD), a heterogeneous form of acute lung injury associated with multi-organ failure and rejection. We hypothesized that early, bedside plasma biomarkers could capture this biological heterogeneity, identifying phenotypes with differential clinical outcomes.

**Methods:** This two-year prospective single-center observational study enrolled 78 bilateral LUTX recipients. With a real-time point-of-care immunoanalyzer we measured IL-1β, IL-2, IL-6, IFN-γ, TNF, CCL-2, IL-15, Ferritin, and D-dimer 36 hours post-reperfusion. Outcome-agnostic Latent Profile Analysis (LPA) was applied to identify bio-signatures. PGD (grade 3) incidence and clinical outcomes were compared across classes.

**Results:** LPA identified three distinct classes, interpreted as biological sub-phenotypes. The Adaptive phenotype (68%) had the lowest inflammatory activation, shortest vasoactive support and Invasive Mechanical Ventilation (IMV) (both 1 day) and ICU stay (3 days), and lowest 72-hours PGD (8%) and Acute Kidney Injury (AKI) (28%) rates. The Hyperinflammatory phenotype (18%) showed the highest IL-6 and Ferritin levels with resolving PGD (69% to 23%, from 6 to 72 hours), but prolonged IMV (6 days), vasoactive support (3 days), ICU stay (7 days), with a high AKI rate (69%). The Coagulopathic phenotype (15%), requiring more intraoperative blood products, exhibited the highest TNF and D-dimer with persistent PGD (36% at 24 and 72 hours), intermediate vasoactive support (2 days), and AKI severity. Six-month rejection-free survival was similar across phenotypes.

**Conclusions:** This preliminary hypothesis-generating study suggests that point-of-care biomarkers after LUTX may identify biological phenotypes with potential clinical relevance. Future studies are needed to confirm such phenotyping.

## Introduction

Lung transplantation (LUTX) is the last therapeutic option for end-stage respiratory failure^1^. The post-operative period after LUTX is characterized by substantial clinical heterogeneity, with recipients experiencing markedly different clinical courses, recovery patterns, and complications. Among these, Primary Graft Dysfunction (PGD) - a form of inflammatory acute lung injury occurring within 72 hours after graft reperfusion - is the most frequent and dreadful, being associated with increased risk of rejection and mortality^2^ and lacking any specific therapy. PGD is currently graded based solely on radiographic criteria and alterations of oxygenation^2^, and its classification does not capture differences in severity^3^, duration^4^, and associated systemic correlates such as hemodynamic failure^5^ and renal dysfunction^6^, which, taken together with respiratory failure, may represent a multi-organ treatable trait.

In other critical illnesses (i.e., acute respiratory distress syndrome – ARDS^7,8^, sepsis^9^), patient subgroups with divergent pathophysiology and potential differential therapeutic responses have been recognized, supporting a precision-medicine treatment approach. Instead, in the context of LUTX, phenotyping remains unexplored, and no study has combined bedside, real-time biomarker assessment with modern statistical methods to detect early molecular signatures predictive of outcomes^10^. Identifying those biological phenotypes may improve PGD risk stratification, potentially enabling early detection of allograft injury and unveiling potential targetable molecular variants^11^.

Based on these premises, we hypothesize that LUTX recipients can be subtyped based on early plasma levels of biomarkers reflecting inflammation, coagulation, and endothelial activation, by means of a real-time, point-of-care (POC) immunoanalyzer.

In this preliminary proof-of-concept prospective observational study, we thus aimed to: 1) measure a panel of biomarkers at the bedside using a POC, real-time biochip-array device; 2) identify distinct biological profiles through outcome-agnostic Latent Profile Analysis (LPA); 3) enrich the biological characterization of these profiles by additional multiplex immunoassays and 4) assess their association with short- and mid-term clinical outcomes.

## Methods

This is a single-center, prospective, observational cohort study performed at an Italian tertiary referral center from June 1^st^, 2023, to June 30^th^, 2025, approved by the Regional Ethical Committee and registered on ClinicalTrials.gov (#NCT06125535). It adheres to the Principles of the Declaration of Istanbul and the Declaration of Helsinki, and follows the STROBE guidelines for observational trials^12^. Written informed consent was obtained from study participants. Adult patients (>18 years) undergoing LUTX were eligible for enrollment. Exclusion criteria were: - re-transplantation; - single LUTX; - combined organ transplantation. The management of LUTX recipients is detailed in the Online Supplement (Additional Methods) and has been previously described^5,13–18^.

### Data Collection

At the time of enlisting for LUTX, the following data were collected: age, gender, body mass index (BMI), comorbidities, and Lung Allocation Score (LAS)^19^. The following intra-operative data were collected: use of Extracorporeal Membrane Oxygenation (ECMO) during LUTX and at the end of surgery; the use of blood components and the relative amount. The following donor data were collected: age and type of donor (i.e., Donor after Brain Death – DBD; Donor after Cardiac Death – DCD); OTO score^20^; warm and cold ischemia times; use of Ex-Vivo Lung Perfusion (EVLP)^21^.

The primary endpoint was a composite of developing PGD, according to the latest guidelines^2^, or death at 24, 48, and 72 hours after first-implanted graft reperfusion. Secondary endpoints included measures of clinical course such as the duration of invasive mechanical ventilation (IMV); 28-day ventilation-free days; 28-day intensive care unit (ICU)-free days; duration of vasoactive support; 28-day vasoactive-free days; and 60-day hospital-free days. Additional clinically observed outcomes included the need for tracheostomy, occurrence of Acute Kidney Injury (AKI) within 7 days^22^ and Acute Kidney Disease (AKD) between 7-90 days^23^; need for post-operative renal replacement therapy (RRT); biopsy-proven acute rejection at 6 months^24^, and survival on July 30th, 2025. All event-free-day outcomes were calculated as composite endpoints combining the specific outcome with death (with deceased patients assigned zero free days).

### Study Procedures

Blood samples were collected from all enrolled patients at two time points: 6 and 36 hours after the first implanted graft reperfusion. Samples were collected in plastic vacuum EDTA containers, centrifuged (1500 G for 15 minutes), and plasma was fresh-frozen and stored at -80°c until analyses.

To detect biological signatures by LPA analysis, after thawing, plasma was assayed for concentrations of the following biomarkers using a point-of-care (POC), real-time, biochip-array device (Multistat, Randox, Coleraine, UK): Interleukin (IL)-1β, IL-2, IL-6, Interferon-gamma (IFN-γ), Tumor Necrosis Factor (TNF), chemokine (C-C motif) ligand-2 (CCL-2), IL-15, Ferritin, and D-dimer. The Multistat device is a fully automated immunoanalyzer that is operable by clinical staff, providing results within a clinically actionable timeframe (< 90 minutes). Although biomarker assays were performed in batch for logistical reasons, the timing of sample collection and the analytical approach closely simulate real-time bedside implementation, with results available within a clinically relevant timeframe for decision-making.

Moreover, to biologically enrich and validate the phenotypic classification derived from POC biomarkers, plasma samples were also analyzed using a Luminex multiplex platform (Bio-Rad, Hercules, California, USA), according to the manufacturer’s instructions. Biomarkers were selected based on findings from a previous scoping review^10^ and included, biomarkers of inflammation (IL-8, Chemokine (C-X-C motif) Ligand 10 (CXCL10), IL-10, Granulocyte-Macrophage Colony-Stimulating Factor (GM-CSF), IFN-α, endothelial dysfunction (Angiopoietin 1 (ANG-1), Angiopoietin 2 (ANG-2), Intercellular Adhesion Molecule 1 (ICAM-1), P-selectin), tissue damage (Tissue Inhibitor of Metalloproteinases-1 (TIMP-1), soluble Receptor for Advanced Glycation End-products (sRAGE), Surfactant Protein D (SP-D)) and coagulation and systemic activation (Procalcitonin, Protein C, soluble-TNF Receptor I (sTNFR1)). The ANG 2/1 ratio (higher values representing pro-inflammatory status) was calculated^25^.

For all the biomarkers, values below the lower limit of quantification were set to the lowest accurately measurable value, whereas values exceeding the upper limit of detection were set to the upper limit of quantification.

### Statistical Analysis

Continuous variables were summarized as median and interquartile range (IQR), while categorical variables were expressed as counts and percentages. Biomarker concentrations were log-transformed and standardized (mean = 0, SD = 1).

To identify latent biological subgroups within a clinically meaningful framework, a Latent Profile Analysis (LPA) was performed exclusively on biomarkers measured at the bedside using the POC real-time biochip-array device (IL-1β, IL-2, IL-6, IFN-γ, TNF, CCL-2, IL-15, ferritin, and D-dimer), obtained 6 and 36 hours after graft reperfusion^26^. Instead, multiplex immunoassays were used only for classification enrichment. Patients identified as outliers for POC biomarkers (based on kurtosis analysis, p < 0.001) were excluded. Models with one to three latent classes were tested, assuming equal variances across classes and no covariances between indicators. Model fit was evaluated using the sample-size–adjusted Bayesian Information Criterion (saBIC), class size (>10 subjects), and entropy (with higher values indicating greater classification certainty). Outcomes were compared using the Bolck–Croon–Hagenaars method to account for assignment uncertainty^27^. Given the exploratory nature of the study, no formal a priori sample size calculation was performed. To mitigate risks associated with small samples, model adequacy was assessed by post hoc diagnostics (entropy and class stability).

Biomarker concentrations at 6 and 36 hours were first compared, regardless of phenotype, using the Wilcoxon signed-rank test. Then, differences in temporal biomarker trajectories among classes were then analyzed using linear mixed-effects models including fixed effects for class, time, and their interaction (class × time), with random intercepts for patients.

All statistical tests were two-sided, and a p-value < 0.05 was considered statistically significant. Analyses were performed using R (version 4.3.2; R Foundation for Statistical Computing, Vienna, Austria) and JMPpro17 (SAS Institute Inc., USA).

## Results

### Study Population and Clinical Outcomes

From June 1^st^, 2023, to June 30^th^, 2025, 97 patients underwent bilateral LUTX at our Institution, and, after excluding 12 patients, 85 met the inclusion criteria for the study (see Figure S1, Online Supplement, Additional Results). In 3 cases, due to logistical constraints, patients could not be recruited in the study. In the remaining 82 recruited patients, 4 had to be excluded due to plasma samples that were lost, corrupted, or destroyed due to processing errors. Thus, 78 patients were included in the final analysis. Table 1 shows the patients’ clinical characteristics. Enrolled patients were predominantly male (59%), with a median age of 56 [45-62] years old. Only 7 patients (8.5%) had non-European origin (3 South American and 4 North African). The most common indication for LUTX enlistment was Interstitial Lung Disease (57%).

**Table 1.**
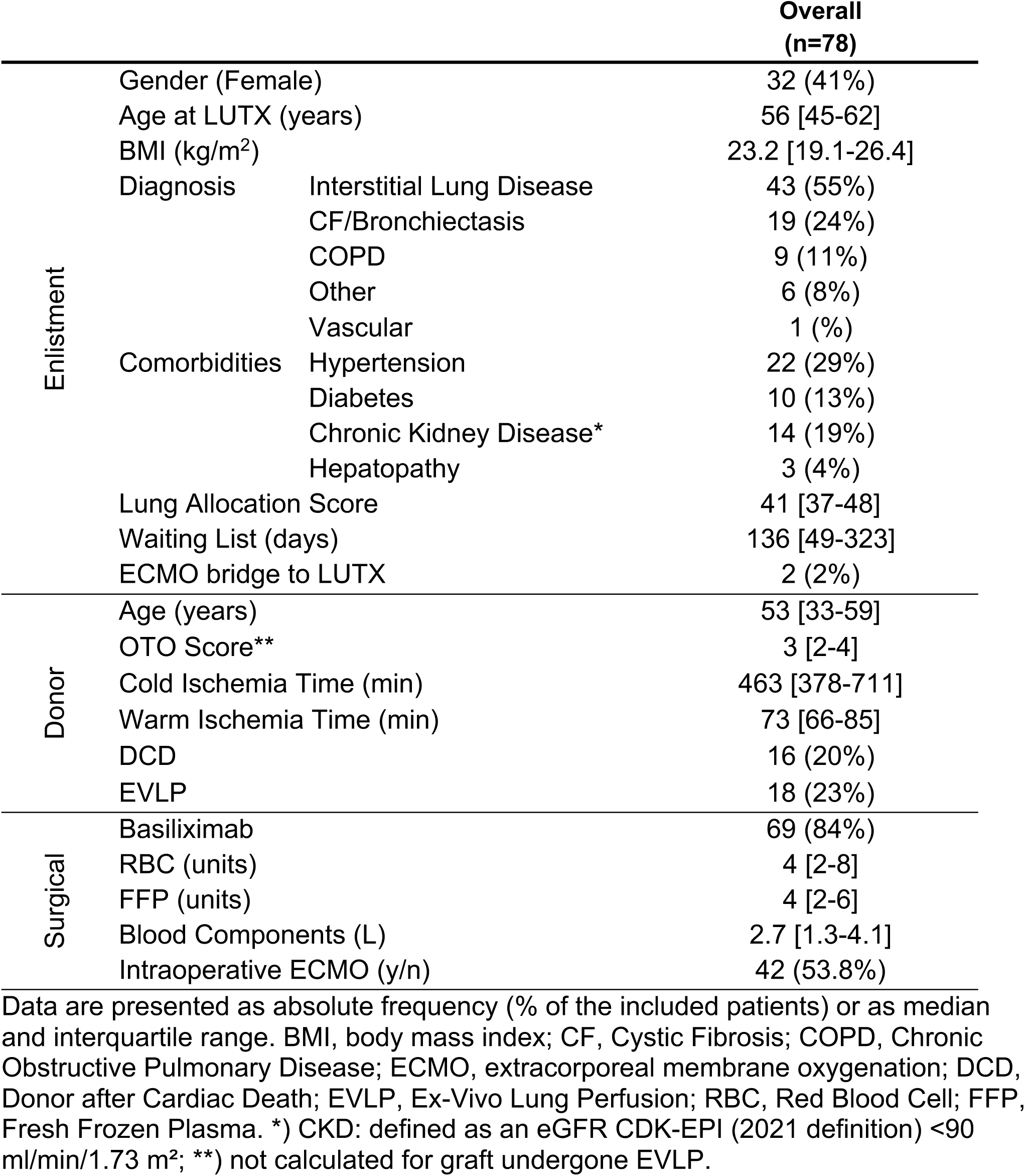
Patients’ characteristics.

### Biomarker Assessment and Temporal Dynamics

Plasma biomarker concentrations in the overall patient population are shown in Table 2. As per the POC device biomarkers, between 6 and 36 hours after reperfusion, IL-6 and CCL-2 decreased significantly, whereas IL-15 and Ferritin showed a significant increase. Among multiplex immunoassays, we observed a statistically significant reduction in biomarkers of inflammation (IL-8, IL-10, GM-CSF), endothelial dysfunction (ICAM-1), and tissue damage (TIMP-1, s-RAGE, and SP-D). In contrast, ANG2 and ANG 2/1 ratio, as well as Protein C, showed a significant temporal increase.

**Table 2.**
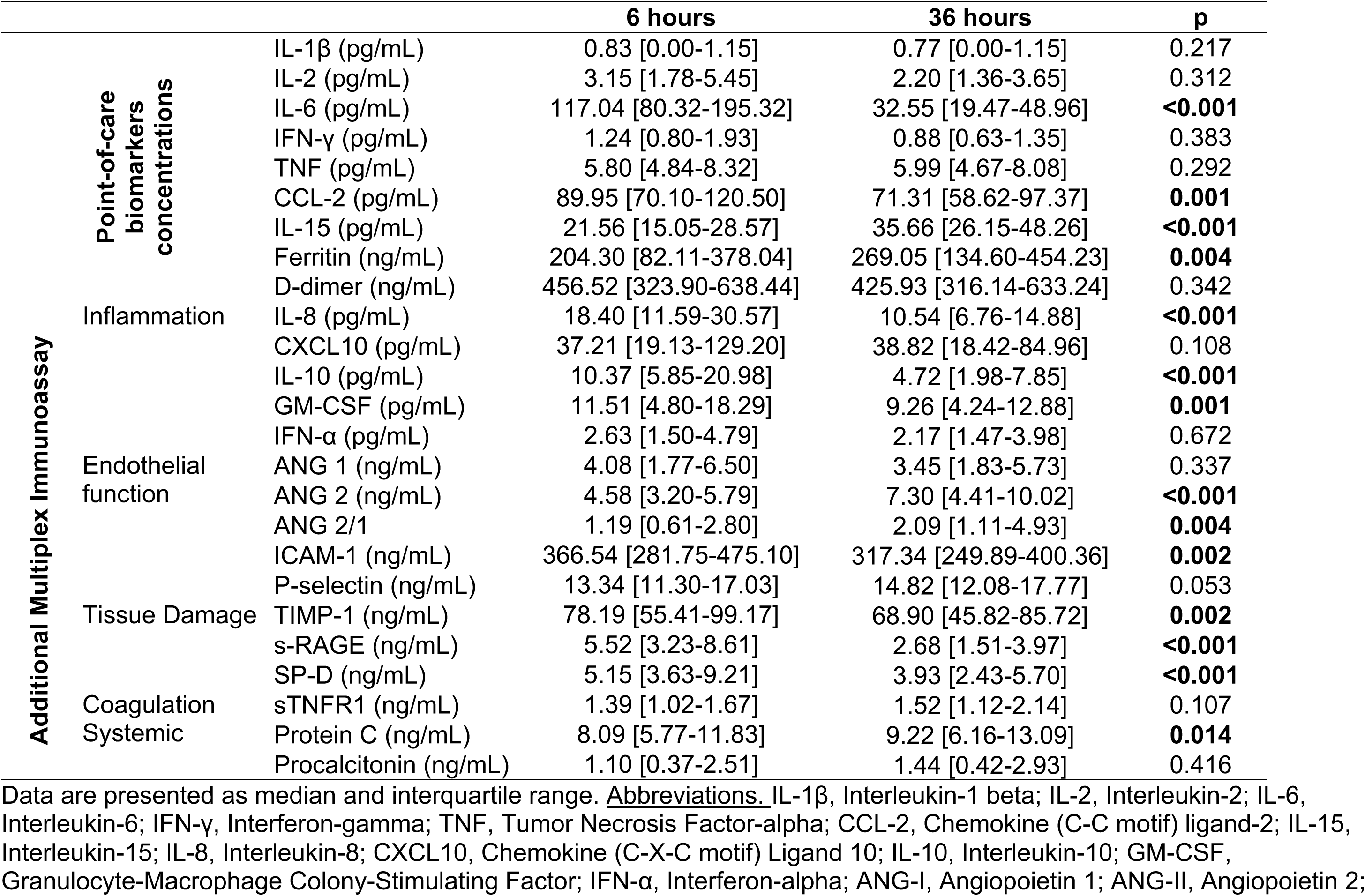
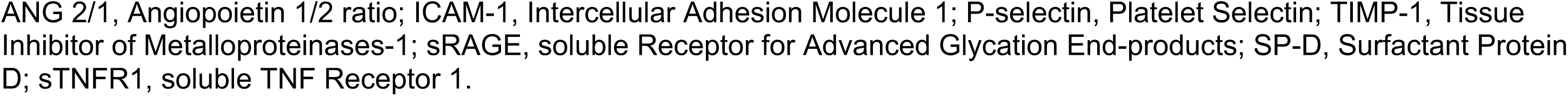
Plasma biomarker concentrations measured at 6 and 36 hours after reperfusion.

### Biological Patterns

Early biological patterns at 6 hours post-transplantation showed no clinically meaningful associations with outcomes and are reported in the Online Supplement (Tables S1, S2, S3). At 36 hours, kurtosis analysis identified 4 patients as statistical outliers (p<0.001), who were excluded from the LPA. The final LPA cohort comprised 74 patients, with complete biomarker measurements. The 3-class solution achieved an entropy of 0.83, whereas the 2-class solution showed poor classification certainty (entropy = 0.65). The saBIC demonstrated progressive improvement with increasing model complexity, decreasing from 1901.7 (1-class) to 1856.6 (3-class solution).

The LPA identified three distinct biological signatures with statistically significant differences across all POC biomarkers, except IL-2 and IFN-γ (Table 3, Figure 1, Panel A and Figure S2). The three subgroups included 50 (68%), 13 (18%), and 11 (15%) patients. The most populated class (68% of patients) comprised patients with the lowest inflammatory profile, and was named the Adaptive phenotype. The second most represented class (18%) had the highest inflammatory burden (particularly IL-6 and ferritin), with the lowest D-dimer, and was named the Hyperinflammatory phenotype. The least represented class (15%) had a relatively high inflammatory burden, and was characterized by elevated D-dimers and TNF, with suppressed IL-1β, and was named the Coagulopathic phenotype.

**Table 3.**
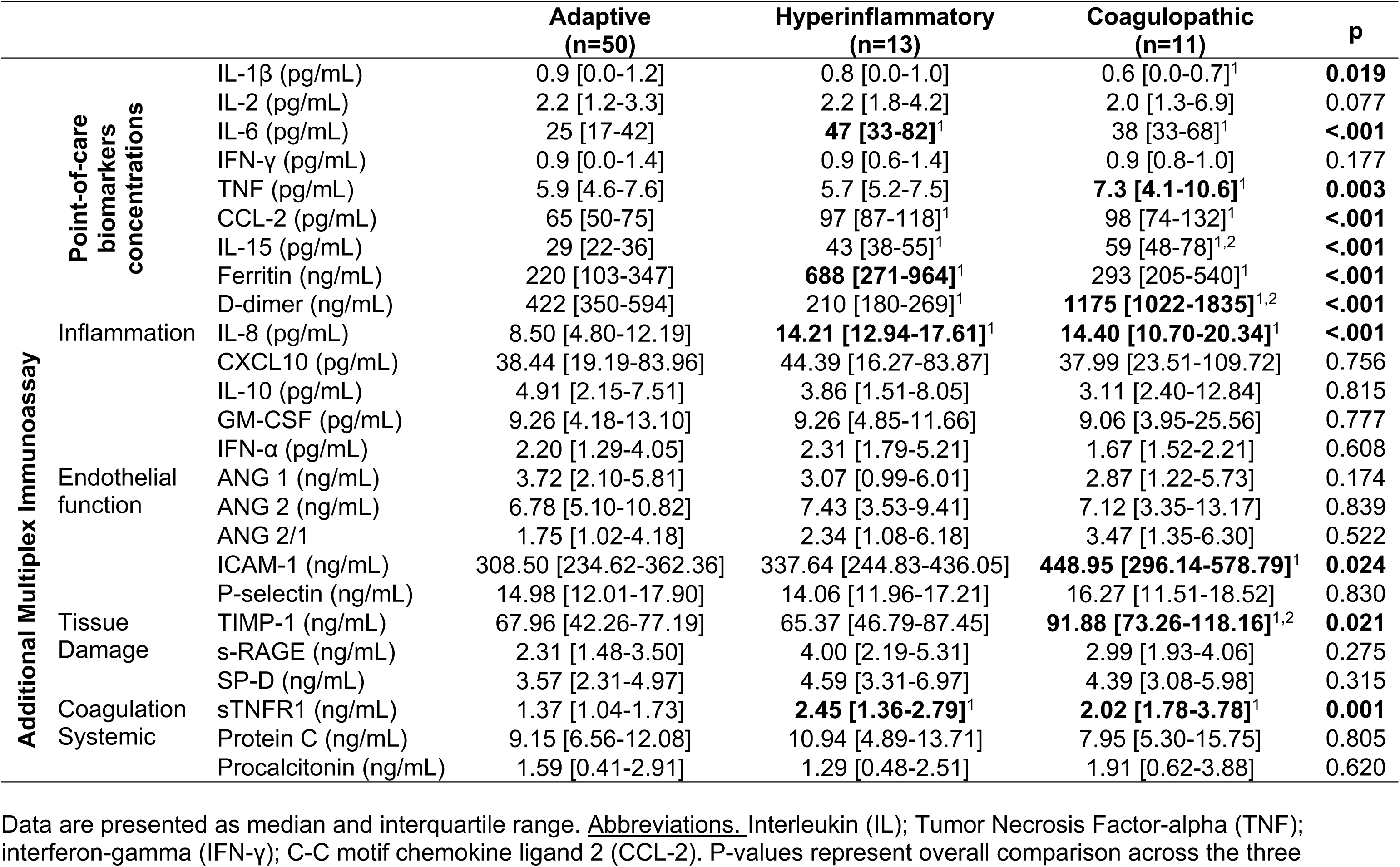
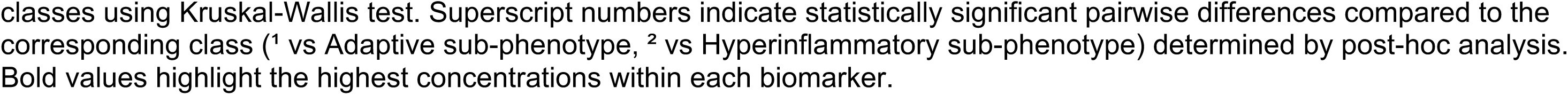
Plasma biomarkers concentrations at 36-hours, stratified by phenotype.

**Figure 1.**
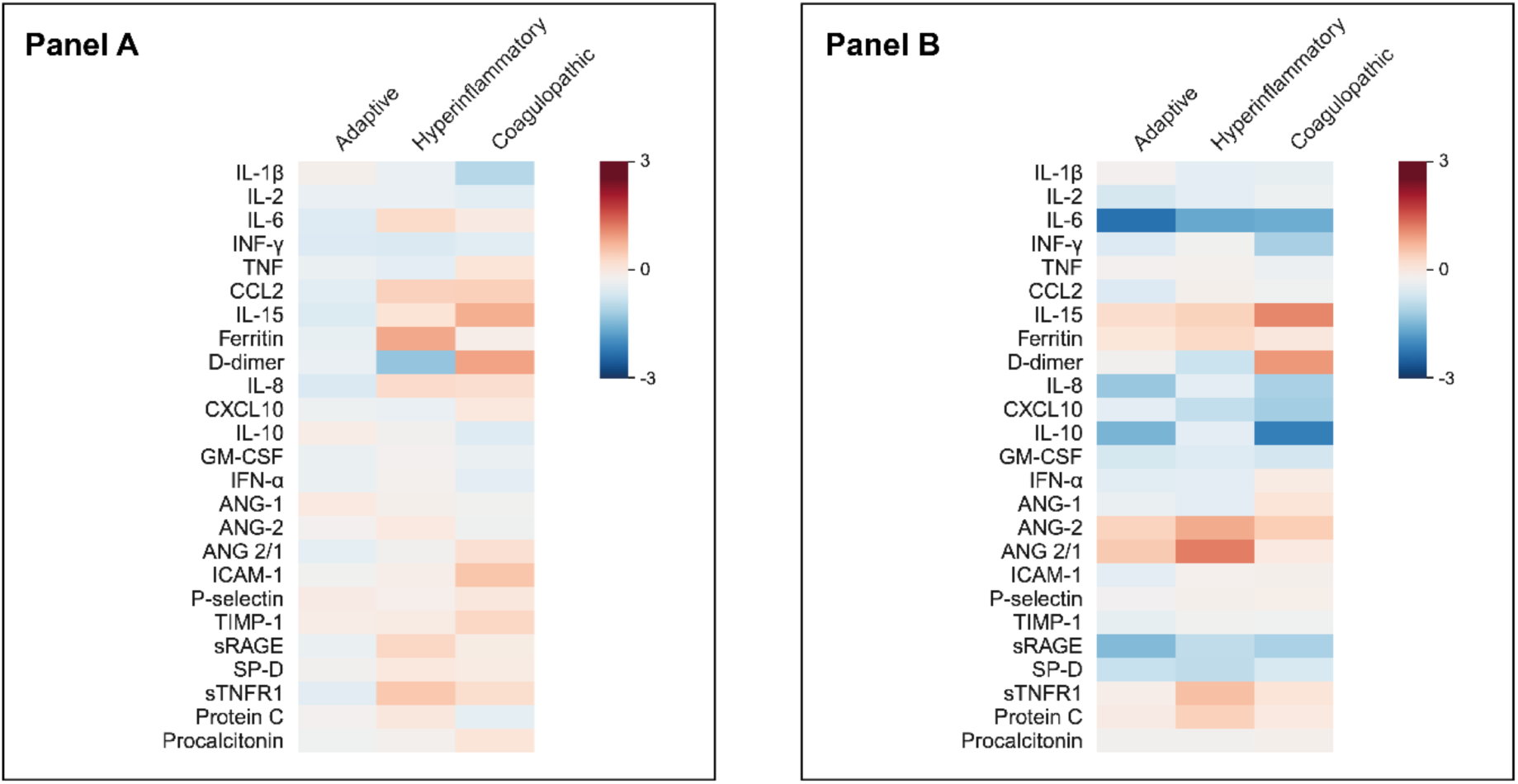
Plasma biomarker concentrations and their temporal trajectories stratified by phenotypes. Panel A. Heatmap of biomarker concentrations stratified by phenotypes. Columns represent phenotypes; rows represent individual biomarkers. Color scale indicates standardized concentrations: red represents higher concentrations and blue represents lower concentrations relative to the cohort mean. Panel B. Heatmap depicting temporal changes in biomarker concentrations from 6 to 36 hours post-LUTX, stratified by phenotype. Color scale represents log₂(fold-change) values: blue indicates decreased concentrations (values <1), white indicates no change (value =1), and red indicates increased concentrations (values > 1). Sample sizes: Adaptive (n=50), Hyperinflammatory (n=13), Coagulopathic (n=11).

After defining phenotypes using bedside POC biomarkers, we further characterized their biological profiles by multiplex immunoassays, see Table 3 and Figure 1, Panel B. The Adaptive phenotype patients had the lowest IL-8 levels and sTNFR1, as compared to both non-adaptive phenotypes. Moreover, the Coagulopathic phenotype had higher ICAM-1 levels compared to the Adaptive phenotype, and higher TIMP-1 compared to both the other classes.

Changes in biomarker concentrations over time (6 to 36 hours) are detailed in Figure 1 Panel B and Table S4. Mixed linear model analysis revealed distinct temporal biomarker trajectories across the three phenotypes.

The Adaptive phenotype showed a resolution of inflammation, with significantly greater reductions in IL-6 (4.8-fold decrease, p=0.035) and ICAM-1 (1.2-fold decrease, p=0.018) compared to the Hyperinflammatory phenotype. Compared with non-adaptive phenotypes, IL-1β and D-dimer were stable (p=0.043 vs. Coagulopathic; p<0.001 vs. non-adaptive phenotypes), while IL-15 showed the smallest increase (1.4-fold, p<0.001 vs.

Coagulopathic). The Adaptive phenotype also showed the large magnitude – albeit non-statistically significant – decreases in sRAGE (2.5-fold), IL-10 (2.6-fold), IL-8 (2.2-fold), and SP-D (1.6-fold decrease).

Contrarily, the Hyperinflammatory and Coagulopathic phenotypes both showed impaired inflammatory resolution. IL-6 reductions were smaller (2.9-fold and 2.8-fold, respectively) than in the Adaptive phenotype, and IL-1β decreased (1.2-fold, p =0.049), contrasting with stable Adaptive levels. IL-15 increased in both but more markedly in Coagulopathic (2.8-fold vs. 1.6-fold, p <0.001).

The Hyperinflammatory phenotype had endothelial and macrophage activation-like biological signatures: ANG-2/ANG-1 ratio increased 2.9-fold, driven by rising ANG-2 (2.2-fold) without ANG-1 compensation, marked sTNFR1 increase (1.9-fold), persistently elevated ferritin, and elevated CCL-2. IL-8 decreased minimally (1.2-fold), the smallest reduction among phenotypes, while D-dimer decreased significantly (1.5-fold, p <0.001). Instead, the Coagulopathic phenotype showed progressive endothelial and coagulation dysfunction, with the large D-dimer increases (2.5-fold, p <0.001), stable ICAM-1 levels (p=0.018), and TIMP-1 remained elevated.

Table 4 shows the enlistment, intraoperative, and donor-related characteristics, stratified by phenotype. Overall, there were no clear differences in clinical features among the three biological phenotypes, except for ILD (more predominant in the Hyperinflammatory class). Clinical outcomes, detailed in Figure 2, Figure 3, and Table 5, varied markedly according to latent biological signatures. In the overall cohort, 11 (14%) patients had PGD grade 3 at 72 hours. The Adaptive phenotype had the most favorable clinical course, characterized by the lowest PGD incidence with rapid resolution (33% to 8% PGD grade 3, from 6 to 72 hours), shorter duration of vasoactive support (1 day) - with most patients off vasopressors by day 3 - and IMV (1 day), lower rates of AKI (28%), shorter ICU and hospital LOS (3 and 23 days, respectively) compared to the other classes.

**Table 4.**
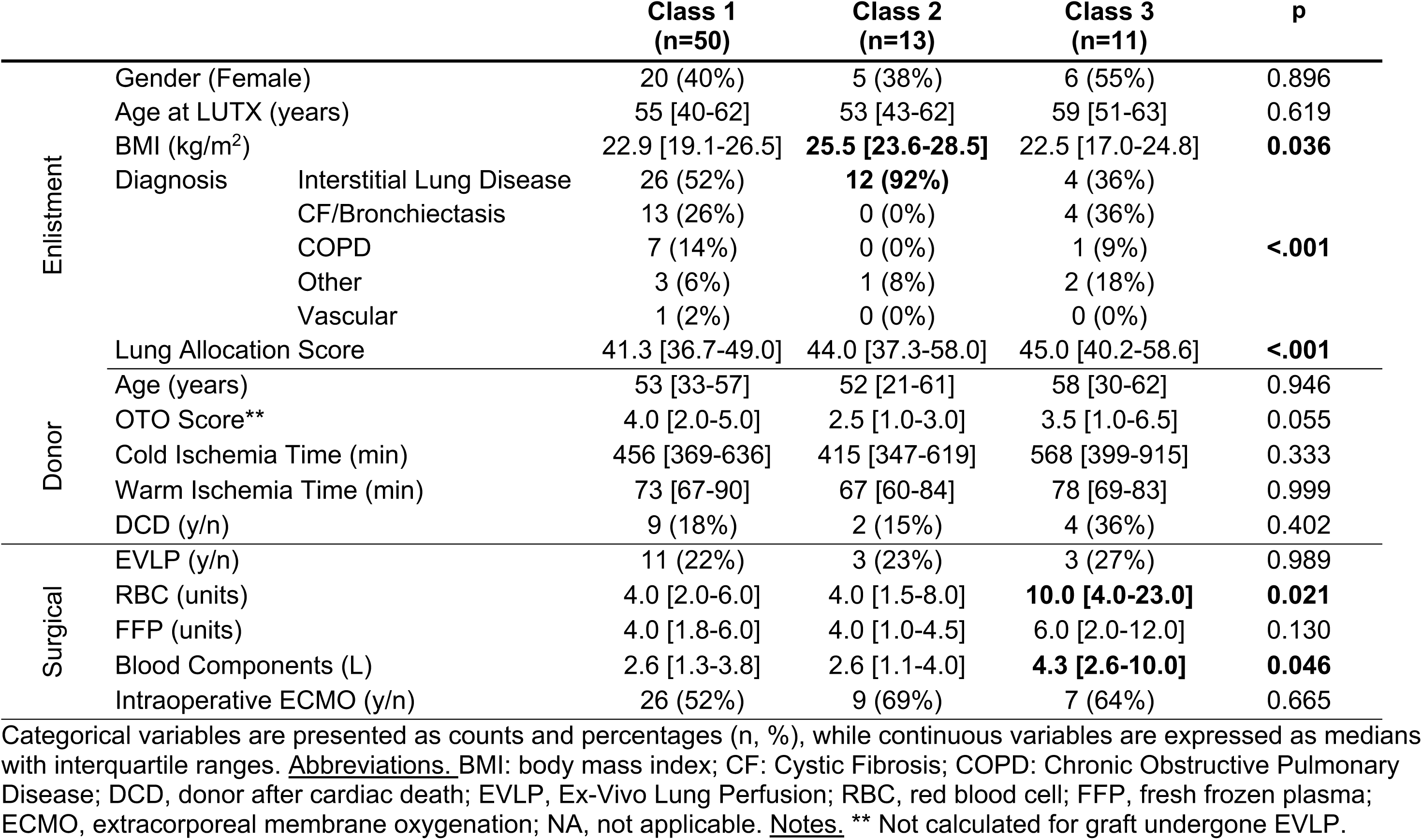
Patients’ characteristics stratified by latent biological classes based on 36-hour point-of-care biomarkers.

**Figure 2.**
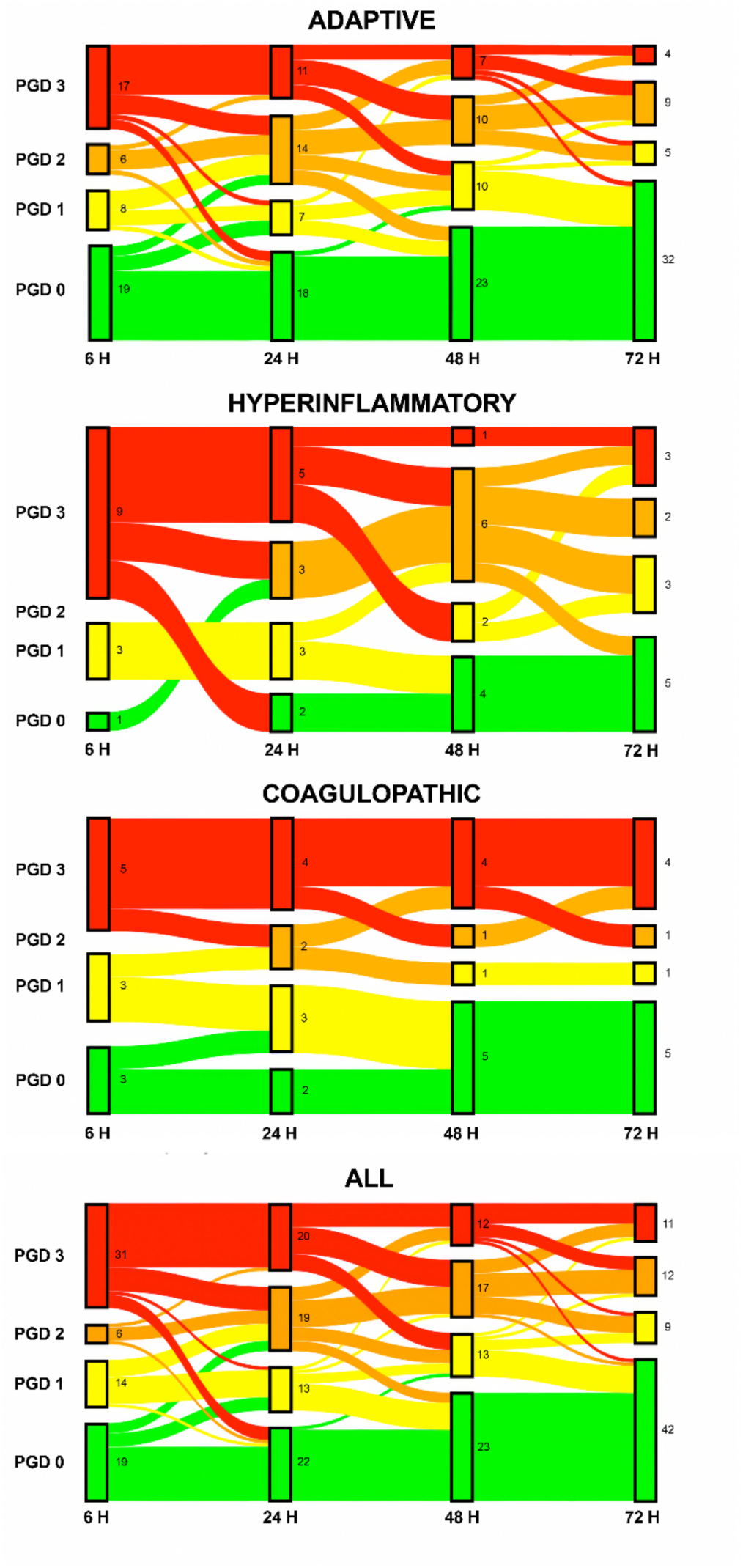
Temporal trajectory of Primary Graft Dysfunction (PGD) in the overall population, and stratified by latent biological classes based on 36-hour point-of-care biomarkers. Sankey diagram illustrating the progression and resolution patterns of PGD grades over the first 72 hours after LUTX. Flow widths are proportional to the number of patients transitioning between PGD grades at each time point (6, 24, 48, and 72 hours). Each horizontal band represents a PGD grade (0, 1, 2, or 3), with connecting flows demonstrating patient transitions between severity grades over time. Upward flows indicate PGD progression (worsening), downward flows represent improvement, and horizontal flows show stability within the same grade. Panel A: overall patients population

**Figure 3.**
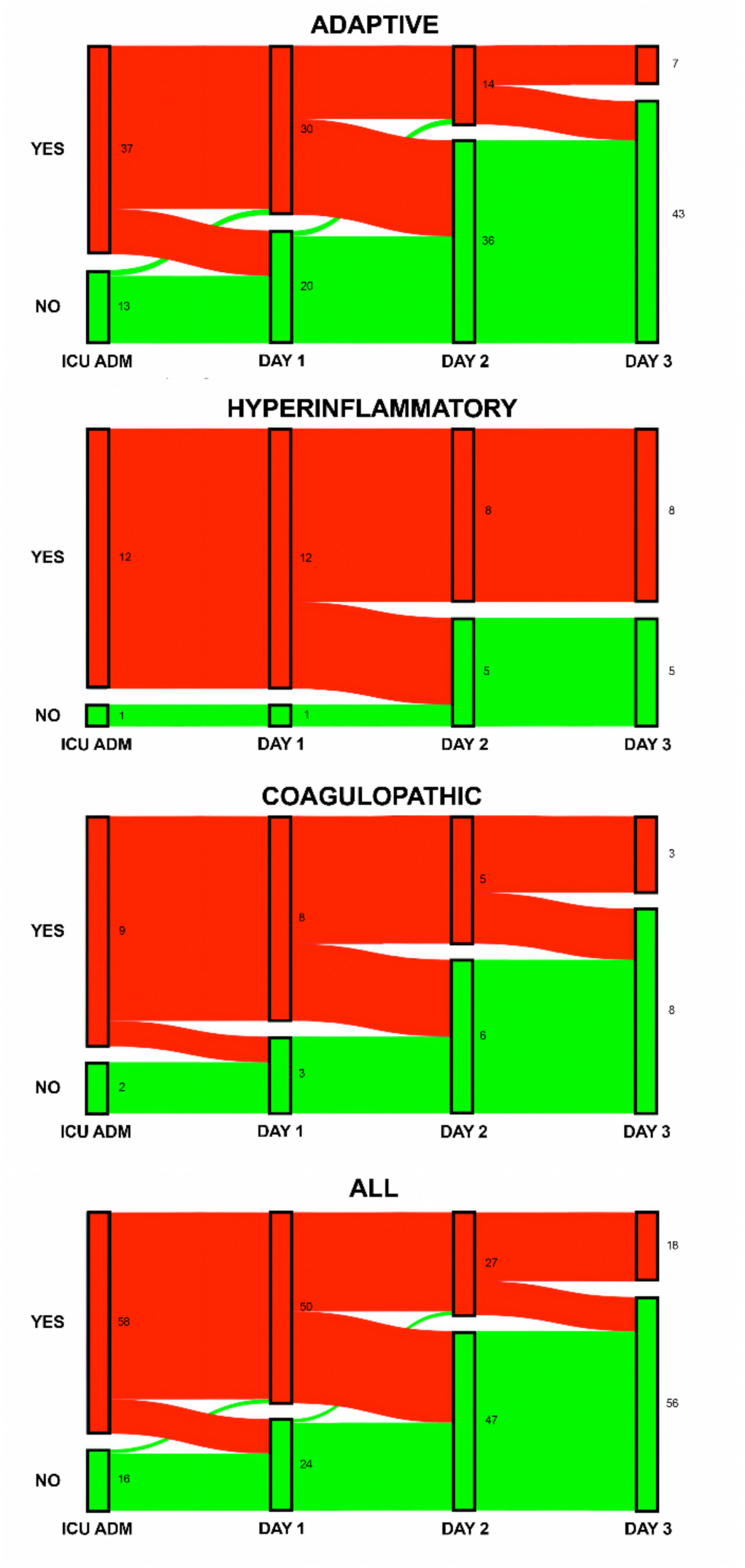
Temporal trajectory of vasoactive needs stratified by latent biological classes based on 36-hour point-of-care biomarkers. Sankey diagram illustrating the progression and resolution patterns of vasoactive need over the first 72 hours after LUTX. Flow widths are proportional to the number of patients transitioning between vasoactive need at each time point (6, 24, 48, and 72 hours). Each horizontal band represents a vasoactive class (yes/no), with connecting flows demonstrating patient transitions between severity grades over time. Upward flows indicate vasoactive introduction (worsening), downward flows represent improvement, and horizontal flows show stability.

**Table 5.**
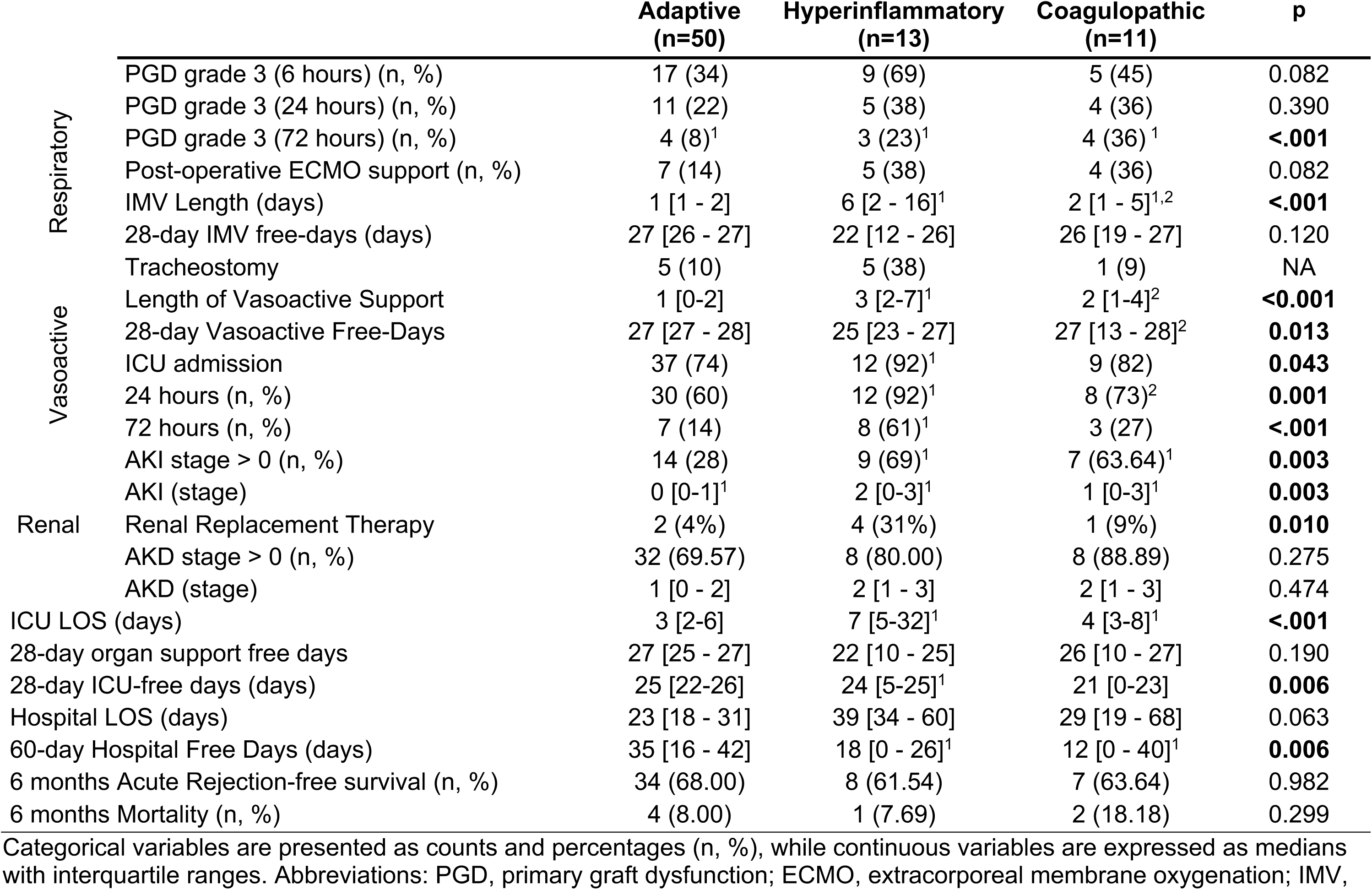
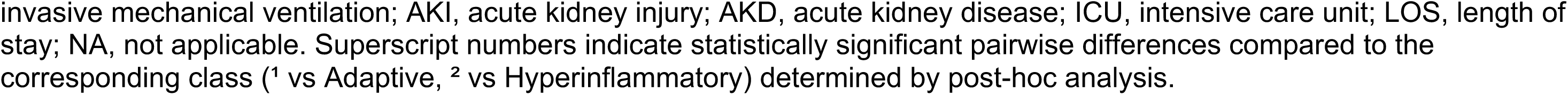
Patients’ outcomes stratified by latent biological classes based on 36-hour point-of-care biomarkers.

In contrast, non-adaptive phenotypes were associated with significantly worse outcomes: higher risk for PGD (> 25% PGD grade 3 at 72 hours), greater organ support requirements, higher AKI rates (>60 %), and prolonged ICU (7 and 4 days, in Hyperinflammatory and Coagulopathic phenotypes, respectively) and hospital LOS (39 and 29 days, respectively).

The Hyperinflammatory and Coagulopathic phenotypes demonstrated divergent temporal evolution in short-term outcomes. Specifically, PGD improved over time in the Hyperinflammatory phenotype (grade 3 PGD decreasing from 69% at 6 hours to 23% at 72 hours), whereas it persisted in the Coagulopathic phenotype (stable at roughly 35%). Early vasoactive support was frequent (92%) in the Hyperinflammatory phenotype, and persisted up to 72 hours (61%), while it was less frequent and resolved in the Coagulopathic phenotype (73% to 27%).

Finally, the Hyperinflammatory required the longest duration of IMV and ICU LOS. Six-month rates of acute rejection and mortality were similar across all biological signatures.

## Discussion

In this preliminary prospective observational study, we identified three distinct biological profiles in LUTX recipients based on bedside-measured inflammatory, tissue damage, and coagulation signatures using an outcome-agnostic latent profile analysis (LPA). Despite similar baseline characteristics, different signatures had differential clinical outcomes and temporal biomolecular trends, suggesting the possibility of real-time phenotyping LUTX recipients for early risk stratification and – eventually – targeted, precision-medicine-based management.

LUTX is the last therapeutic option for end-stage respiratory failure. Despite recent advances in surgical techniques and immunosuppressive regimens, LUTX is frequently burdened by PGD, a heterogeneous form of inflammatory acute respiratory failure accompanied by multi-organ dysfunction^17,28^, and, in the long term, associated with rejection^2^. In this scenario, identifying biological signatures may improve risk stratification and detection of early LUTX phenotypes that represent treatable traits.

In a previous structured scoping review^10^, we documented a substantial knowledge gap on this topic. Although few previous studies have measured multiple biomarkers in LUTX recipients^29–31^, none have attempted to analyze them using an outcome-agnostic clustering analysis. Unlike previous studies that stratified patients based on predetermined clinical outcomes (e.g., PGD vs. non-PGD, AKI vs. non-AKI), in this work, LPA allowed the biological data itself to define patient subgroups without preconceived syndromic classifications. This approach has been shown to be valid in other critical syndromes, such as ARDS^7,8^ and sepsis^9^, documenting the existence of distinct phenotypes with potential differential treatment effect.

For biomarker selection in phenotyping, we chose the RANDOX POC platform for several strategic reasons. First, this POC device provides results within a clinically meaningful 1-hour timeframe, potentially enabling real-time biological phenotyping at the bedside. To our knowledge, use of such technology has never been described in transplant recipients. Second, among the available POC biomarker kits, we selected the array offering the broadest spectrum of biologically relevant markers. While this panel was not designed explicitly for LUTX, it includes highly relevant pathophysiological domains, which were separately assessed in several LUTX cohorts: pro-inflammatory cytokines (IL-1β^32^, IL-6^33–35^, TNF^36,37^), chemokines mediating neutrophil recruitment (CCL-2^36^), immune regulatory markers associated with T-cell responses (IL-2^38^, IL-15, IFN-γ^31^), acute phase reactants (ferritin), and coagulation markers (D-dimer). IL-2 may be of particular interest in LUTX recipients, given that one of the most commonly used immunosuppressants (i.e., basiliximab) is a potent inhibitor of IL-2 activity^39^. To further enrich the phenotyping analysis, we complemented the POC measurements with additional biomarkers measured by Luminex-based multiplex immunoassays, with biomarker selection informed by our previous scoping review and existing literature.

We identified three distinct biological phenotypes based on 36-hour biomarker profiles, each characterized by unique biological signatures that correlated with significantly different clinical outcomes. Unlike conventional PGD grading, these biomarker-defined phenotypes captured a much broader multi-organ clinical complexity of LUTX recipients. Notably, 6-hour biomarker profiling did not result in the discrimination of clinically different cohorts, suggesting that biomarker differentiation becomes apparent only after sufficient time following ischemia–reperfusion injury.

The most represented subgroup (around 70%) had low inflammatory mediators and an inflammatory resolution pattern, suggesting an adaptive transition from ischemia-reperfusion injury to tissue repair and endothelial recovery, leading to overall good outcomes: lowest PGD incidence, minimal vasoactive support, lowest AKI rates, shortest IMV, ICU, and hospital LOS. For these reasons, this class was defined as Adaptive phenotype. The rest of the patients shared some common pro-inflammatory patterns, but among those non-adaptive patients, two distinct subgroups were recognized.

Around 20% had high IL-6 and ferritin levels, and the lowest D-dimer levels. These patients had early severe PGD that improved over time, but still required prolonged vasoactive support, with frequent and severe AKI, leading to prolonged IMV, RRT requirements, ICU, and hospital stays. The temporal evolution analysis revealed persistent inflammatory activation with macrophage-activation-like patterns and endothelial dysregulation, and thus was called the Hyperinflammatory phenotype. The least represented cohort (15%) was characterized by the highest TNF levels, IL-15, and D-dimers, persistent endothelial activation by ICAM-1 and TIMP-1, suggesting a cascade of inflammation-driven endothelial injury leading to intravascular coagulation activation.

These patients required significantly more intraoperative blood products (10 vs. 4 RBC units), suggesting consumption coagulopathy in the setting of severe inflammation. For this reason, this cohort was named the Coagulopathic phenotype. Clinically, they showed delayed persistent PGD with intermediate vasoactive support and moderate AKI rates, which overall led to prolonged ICU and hospital LOS.

The complementary analyses obtained by Luminex-based multiplex analyses documented that both Hyperinflammatory and Coagulopathic phenotypes shared elevated IL-8 and sTNFR1, biomarkers characteristic of the hyperinflammatory ARDS phenotype^40^. These observations suggest convergent inflammatory pathways despite different clinical contexts. Notably, traditional ARDS phenotyping variables such as vasopressor requirements and bicarbonate levels would not have been useful for phenotype characterization in LUTX, since vasopressor use in the immediate post-transplant period reflects specific intraoperative hemodynamic alterations (e.g., blood loss, effect of sedative), while bicarbonate levels are influenced by massive intraoperative fluid administration. Additionally, ARDS phenotyping is applied *after* ARDS diagnosis, whereas our approach represents preemptive phenotyping before PGD development.

Regardless of phenotype, IL-2 and IL-15 demonstrated contrasting patterns, providing insights for personalized, real-time monitored immunosuppression. Despite sharing receptor components and JAK-STAT signaling pathways, they differ in their α-chain expression (i.e., IL-2Rα (CD25) on activated T/B cells vs. IL-15Rα on monocytes/dendritic cells), and thus are critically different in their susceptibility to basiliximab (an anti-CD25 monoclonal antibody) immunosuppression. Our findings reflected this distinction: IL-2 was consistently suppressed, while IL-15 demonstrated progressive elevations, suggesting that T-cell responses are controlled and that the monocyte-dendritic cell axis escapes the current immunosuppressive strategies.

Beyond the universal IL-2/IL-15 pathway, each biological sub-phenotype may present distinct treatable traits that could guide precision management approaches. The Adaptive Phenotype, characterized by successful inflammatory resolution and fast recovery, may benefit from Enhanced Recovery After Surgery (ERAS) protocols, such as fast-track extubation and mobilization. The Hyperinflammatory Phenotype may be targeted with tocilizumab (IL-6 receptor antagonist), potentially mitigating the prolonged inflammatory cascade. In fact, a randomized controlled trial (NCT06033196) wherein LUTX recipients are randomized to non-targeted IL-6 blockade is ongoing. Treating just patients with the Hyperinflammatory Phenotype with tocilizumab would allow precision-medicine treatment, limiting harm (i.e., infectious complications) to non-susceptible traits. The Coagulopathic Phenotype may instead benefit from targeted post-operative hemostatic support with antifibrinolytic agents or factor concentrates, or an immunosuppressant regimen based on anti-ICAM^41^ agents.

These phenotype-specific therapeutic targets may represent a paradigm shift from uniform post-transplant management toward personalized, biologically-guided interventions, but remain hypothetical, since our study has several limitations. First, the sample size is insufficient for a robust LPA. While our entropy values indicated good classification certainty, more populated cohorts (i.e., > 300 patients) are necessary to confirm the phenotypic structure. Second, as a single-center study, the generalizability of our findings is limited. Third, our cohort had limited ethnic diversity, which limits the generalizability of our findings to more diverse populations. Fourth, we lacked an independent validation cohort to confirm the reproducibility of the biological phenotypes. Fifth, our study lacked transcriptomic data that could provide deeper mechanistic insights into the pathophysiological pathways underlying each biological sub-phenotype. To address these limitations, a prospective multicenter observational study is currently ongoing, which aims to validate these biological phenotypes in a larger, more diverse patient population across multiple transplant centers and explore their association with long-term clinical outcomes. In conclusion, this preliminary study suggests that point-of-care biomarker assessment combined with outcome-agnostic latent profile analysis may identify biologically relevant phenotypes in lung transplant recipients that appear to correlate with short-term clinical outcomes. While these initial findings require validation in larger, more diverse cohorts, they provide a proof-of-concept for potential precision medicine approaches in lung transplantation that could inform future risk stratification strategies and targeted therapeutic interventions.

## Data Availability

All data produced in the present study are available upon reasonable request to the authors

## List of non-standard abbreviations

AKD: Acute Kidney Disease
AKI: Acute Kidney Injury
ANG-I: Angiopoietin I
ANG-II: Angiopoietin II
BMI: Body Mass Index
CCL-2: Chemokine (C-C motif) ligand-2
CXCL10: Chemokine (C-X-C motif) Ligand 10
DBD: Donor after Brain Death
DCD: Donor after Cardiac Death
ECMO: Extracorporeal Membrane Oxygenation
ERAS: Enhanced Recovery After Surgery
EVLP: Ex-Vivo Lung Perfusion
GM-CSF: Granulocyte-Macrophage Colony-Stimulating Factor
ICAM-1: Intercellular Adhesion Molecule 1
ICU: Intensive Care Unit
IFN-α: Interferon-gamma
IFN-γ: Interferon-gamma
ILD: Interstitial Lung Disease
IMV: Invasive Mechanical Ventilation
LAS: Lung Allocation Score
LCA: Latent Class Analysis
LOS: Length of Stay
LPA: Latent Profile Analysis
LUTX: Lung Transplantation
OTO: Organ Transport Organization
PGD: Primary Graft Dysfunction
POC: Point-of-Care
RRT: Renal Replacement Therapy
saBIC: Sample size adjusted Bayesian Information Criterion
SP-D: Surfactant Protein D
s-RAGE: Soluble Receptor for Advanced Glycation End-products
sTNFR1: Soluble Tumor Necrosis Factor Receptor I
TIMP-1: Tissue Inhibitor of Metalloproteinases-1
TNF: Tumor Necrosis Factor-alpha

## Online Supplement

### Additional Methods

#### Lung Transplantation Management

##### LUTX Team

Lung transplantation (LUTX) team at Fondazione IRCCS Ca’ Granda - Ospedale Maggiore Policlinico is composed of two thoracic surgeons, one thoracic surgery fellows, a cardiac surgeon, two certified anesthesiologists, one thoracic anesthesia fellow, and one anesthesia fellow, a perfusionist, an operatory-room nurse and one surgical-nurse.

##### LUTX Anesthetic preparation

After routine monitor as per ASA guidelines, induction of general anesthesia with fentanyl (1-2 mcg/kg), midazolam (1-2 mg) and propofol (1-3 mg/kg), and muscle paralysis with rocuronium (0.6 mg/kg) is achieved. Patients are intubated with large size single-lumen tube and undergo aggressive bronchoscopic toilette. Then, the single-lumen tube is substituted, and lung isolation is achieved with appropriately sized left-sided double-lumen endotracheal tube under bronchoscopic guidance. General balanced anesthesia is maintained with fentanyl (1-2 mcg/kg/hr) and sevoflurane (end-tidal concentration 0.5%-1.0%), and muscle paralysis with sequential rocuronium boluses (0.15 mg/kg). Patients are monitored with invasive right radial artery cannulation, central venous catheterization of the right internal jugular vein, and oximetric pulmonary artery catheterization capable of cardiac output monitoring by means of thermodilution technique. Trans-esophageal echocardiography is implemented pending the anesthesiologist preferences. A large bore (i.e., 7 Fr) catheter is introduced in the right antecubital vein and connected to a custom made rapid infusion system, capable of the infusion of up to 500 mL/min. Throughout the procedure, patients are mechanically ventilated in volume control mode, with FiO_2_ to maintain SpO_2_> 90%, and minute ventilation to limit hypercapnia and acidosis. Antibiotic prophylaxis is provided following the indication of infectious disease specialists and pre-operative airways cultures.

##### LUTX Surgical procedure

Patients are treated with sequential bilateral LUTX, using bilateral anterolateral thoracotomy with a transverse sternotomy or two anterior thoracotomies, pending the anatomical characteristics of the patients. After lysis of pleural and mediastinal adhesions, the vascular structures and bronchi are isolated. Then the less perfused lung (as per pulmonary perfusion scintigraphy) is disconnected from the mechanical ventilation and allowed to deflate. The pulmonary artery is cross-clamped, and the first cross-clamping 10 minutes test is performed: the surgical procedure is halted, hemodynamics are strictly monitored,and blood gas analyses are obtained every 2 minutes.

The veins, artery, andbronchus are sequentially resected. The graft is positioned in the thoracic cavity, and the bronchial, arterial and venous anastomoses are created. The lung is thoroughly de-aired before vascular unclamping to avoid systemic air emboli; the first lung graft is slowly re-perfused and connected to another mechanical ventilator. The graft is initially ventilated in pressure control mode with FiO_2_ 21%, PEEP of 10 cmH_2_O, RR of 4 bpm and plateau pressure of 25 cmH_2_O. A recruitment maneuver is applied to obtain complete lung inflation. Progressively, ventilation and oxygenation of the graft are increased to allow contralateral lung separation from ventilation. Particular attention is paid in limiting 1) FiO_2_ (i.e., < 50%); 2) driving pressures (i.e., < 15 cmH2O) and 3) de-recruitment (i.e., PEEP > 10 cmH_2_O) of the implanted graft. Then, contralateral native lung ventilation is interrupted, and the second pulmonary artery cross-clamping test is performed. Pneumonectomy of the second native lung and implantation of the second graft follows the procedure above. Finally, hemostasis is achieved, bilateral pleural drainage positioned, the thorax closed and the patient is transferred to the intensive care unit for follow-up.

##### ECMO indication and pulmonary artery cross-clamping test

The pulmonary artery cross-clamping test is performed to simulate the hemodynamic conditions occurring during pneumonectomy in a controlled -and reversible-fashion, allowing the anesthesiologist to optimize hemodynamics and ventilation and eventually the cardiac surgeon to implement central veno-arterial ECMO in a semi-elective condition. During the first pulmonary artery cross-clamping test (while the native lung is ventilated and perfused), whether 1) pulmonary hypertension (i.e., systolic pulmonary artery pressure > 80 mmHg, 2) increase in PAPs > 50 mmHg associated with systemic hypotension (i.e., systolic arterial pressure < 60 mmHg) resistant to inotropic support; 3) major cardiac arrhythmias; 4) hypoxia (i.e., PaO_2_< 60 mmHg, despite increasing FiO_2_ up to 100% and optimizing PEEP); 5) respiratory acidosis (i.e., pH < 7.25 despite increasing minute ventilation) the test is interrupted and ECMO implemented.

During the second pulmonary-artery cross-clamping (while the graft is ventilated and perfused), our policy is more protective towards hyperoxia and ventilator-induced lung injury. Thus, during the second test FiO_2_ is not increased > 50% and driving pressure is not increased> 15 cmH_2_O. If one of the mentioned conditions occur, ECMO is implemented.

During all the surgical procedure, intractable hypoxemia, acidosis, and hemodynamic failure may occur at any given moment, but 1) single lung ventilation, 2) cross-clamping tests and 3) reperfusion of the grafts are the most critical. Whether during the procure 1) pulmonary hypertension (i.e., systolic pulmonary artery pressure > 80 mmHg, 2) increase in PAPs > 50 mmHg associated with systemic hypotension (i.e., systolic arterial pressure < 60 mmHg) resistant to inotropic support; 3) major cardiac arrhythmias; 4) hypoxia (i.e., PaO_2_< 60 mmHg, despite increasing FiO_2_ up to 100% to the native lung or 50% to the grafts and optimizing PEEP); 5) respiratory acidosis (i.e., pH < 7.25 despite increasing minute ventilation, while guaranteed driving pressure to the grafts < 15 cmH_2_O); the procedure is briefly halted, and ECMO implemented.

Our approach to intraoperative extracorporeal life support consists of central veno-arterial ECMO. After providing unfractionated heparin (i.e., 5000 UI) and eventual further boluses to achieve an aPTT>40 seconds, the ascending aorta and right atrium are cannulated. Blood is drained via a centrifugal pump directly to a polypropylene membrane lung where blood is oxygenated, decarboxylated, warmed and then directed to the central venous circulation. Initially, blood flow is set to achieve around 50% of the patient’s cardiac output, gas flow to maintain normocapnia and fraction of oxygen in the sweep gas flow to maintain SpO_2_> 95%. The extracorporeal circuit setting is dynamically modified during the procedure, pending the different surgical and anesthetic requirements and mean arterial pressure is maintained > 60 mmHg by increases in extracorporeal blood flow, but complete blood drainage and emptying of the heart is avoided, and the opening of the aortic valve is always guaranteed.

At the end of the surgical procedure, prior to chest closure, patients undergoing VA-ECMO undergo a progressive de-escalation of extracorporeal support, consisting of: 1) reduction of blood flows down to 1 L/min; 2) reduction of extracorporeal FiO2 down to 21%. Hemodynamics are monitored and blood gas analysis are collected serially, to verify the patient do not suffer from: 1) hemodynamic failure/right ventricle failure; 2) hypoxemia; 3) acidosis and hypercapnia and/or need to ventilate the patients with tidal volume > 6 mL/kg or driving pressure > 14 cmH2O. Whether hemodynamic failure is observed, central VA-ECMO is converted to peripheral VA support (i.e., femoral cannulations). Whether hypoxemia and/or hypercapnia is observed, VV-ECMO (i.e., femoral-femoral cannulation) is instituted and prolonged in the post-operatory period. No predefined standard management of blood components is applied, but patient-tailored transfusion management is carried out following blood gas analyses, point of care (POC) PT/aPTT tests, and thromboelastography, as per national guidelines.

##### Immunosuppressive therapy

All recipients received induction therapy corticosteroids (methylprednisolone 1000 mg) at graft reperfusion. Moreover, patients received basiliximab (20 mg intravenously on postoperative days 0 and 4), except in the case of a previous history of neoplastic disease. Maintenance immunosuppression consisted of a triple-drug regimen including a calcineurin inhibitor (tacrolimus, Prograf®, oral formulation, twice a day), an antiproliferative agent (i.e., azathioprine), and corticosteroids. Tacrolimus drug dosing and serum level monitoring followed standard postoperative transplant practice (i.e., tacrolimus through level: 10-15 ηg/mL first 3 months, 8-12 ηg/mL up to 6 months).

##### Post-operative Management

We follow generally accepted guidelines for the post-operative management of LUTX recipients.

Protective mechanical ventilation is guaranteed. We limit tidal volume to 6 mL/kg of donor-predicted body weight and plateau pressure < 30 cmH_2_O. We use the lowest inspired oxygen fraction capable of providing an arterial oxygen saturation > 90%. Positive end-expiratory pressure (PEEP) is set following a decremental PEEP trial, with the usual initial PEEP being >= 10 cmH2O. Higher levels of PEEP are utilized in case of plasmorrhea and/or respiratory failure. Possible step-up therapies employed in case of primary graft dysfunction are prolonging sedation, introducing neuromuscular blockade, and ECMO, as previously mentioned. We pursue early weaning from mechanical ventilation, and thus the standard non-complicated patient is usually rapidly weaned from mechanical ventilation and extubated in the first 24 hours after graft reperfusion employing elective noninvasive ventilation. A dedicated respiratory therapist is available 12 hours/24 hours, 7 days/7 days to tailor the management of noninvasive support. Hemodynamic management is guided by lactate, urinary output, invasive cardiac output monitoring, pulmonary artery pressure, wedge pressure, and mixed venous saturation measurement through an elective pulmonary artery catheter. We do not employ a specific hemodynamic protocol but follow the following general rules of thumb: a restrictive fluid strategy to maintain euvolemia, high-threshold red blood cell transfusions (i.e., > 9 gr/L), and vasopressors are used to guarantee a mean arterial pressure in the 65-75 mmHg range and cardiac index (i.e., 2.2-2.5 L/min/m^2^). Antimicrobial therapy is tailored to patients’ and donors’ characteristics. All patients receive prophylaxis for opportunistic infections: fungal (i.e., voriconazole), pneumocystis jirovecii (i.e., trimethoprim/sulfamethoxazole), and viral (i.e., ganciclovir). All patients receive surgical site infection prophylaxis for G-strains (i.e., cefepime for 72 hours) and G+ strains (i.e., vancomycin, one-shot). Cefepime is interrupted at 72 hours whether the donors’ respiratory cultures are negative. Colonized patients continue their antibiotic treatment throughout the perioperative period. Strict infection surveillance is carried out during the post-operative period, by means of deep respiratory cultures collected at 24 hours, 5 days, and 15 days after LUTX and respiratory, anal, and perineal swabs for multi-drug resistant bacteria.

##### AKI and AKD definition

Serum Creatinine (sCr) was measured and collected at the following timepoints: enlistment, before LUTX (i.e., 6 hours before surgery commencement); each day during the ICU and hospital stay; at any ambulatory follow-up within 90 days of LUTX. Notably, the sCr measured 6 hours before surgery was considered baseline and utilized for diagnosis of renal dysfunction. At the same timepoints, the need for RRT was assessed. UO was assessed daily during ICU and hospital stay, up to the 7th post-operative day. According to KDIGO criteria for AKI and ADQI16 workgroup classification for AKD patients were classified as having: a) post-operative AKI whether sCr increased ≥ 0.3 mg/dL at 48 hours, or in the first 7 days after LUTX sCr increased >1.5 times compared to pre-operative (stage 1, stage 2, and stage 3 whether sCr increased 1.5-1.9, 2-2.9, and >3 times, respectively) or having a UO <0.5 mL/kg/hr for 6-12 hours, < 0.5 mL/kg/hr for >12 hours, <0.3 mL/kg/hr for ≥24 hours or anuria for ≥12 hours; b) post-operative AKD whether sCr increased 1.5-1.9, 2-2.9, and >3 times (or need for dialysis) between 7 and 90 days after LT.

## Authors’ contributions

VS conceived the study; VS, GT, VV, DC, LR collected clinical, respiratory function data and data regarding pre, intra, and post-operative management; LMC, VB performed patients’ follow-up and collected follow-up data; VS, FB performed data analyses; LT, MCS, ET performed Luminex analyses and interpreted cytokine measurements; VS, SMC, and FM drafted the original version of the manuscript; LB, LR, FB, MN and GG revised the manuscript critically; all authors approved the final version of the manuscript.

## Additional results

**Figure S1.**
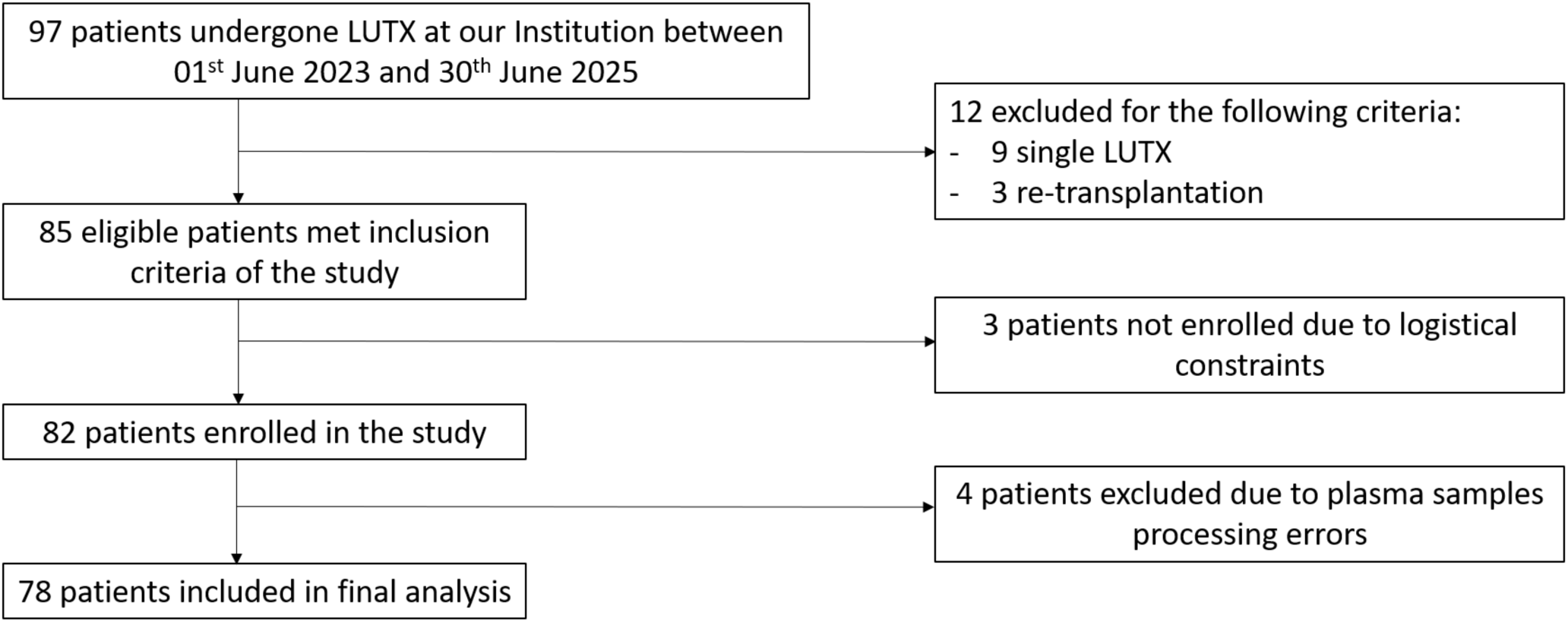
Patients enrollment flowchart.

**Table S1.**
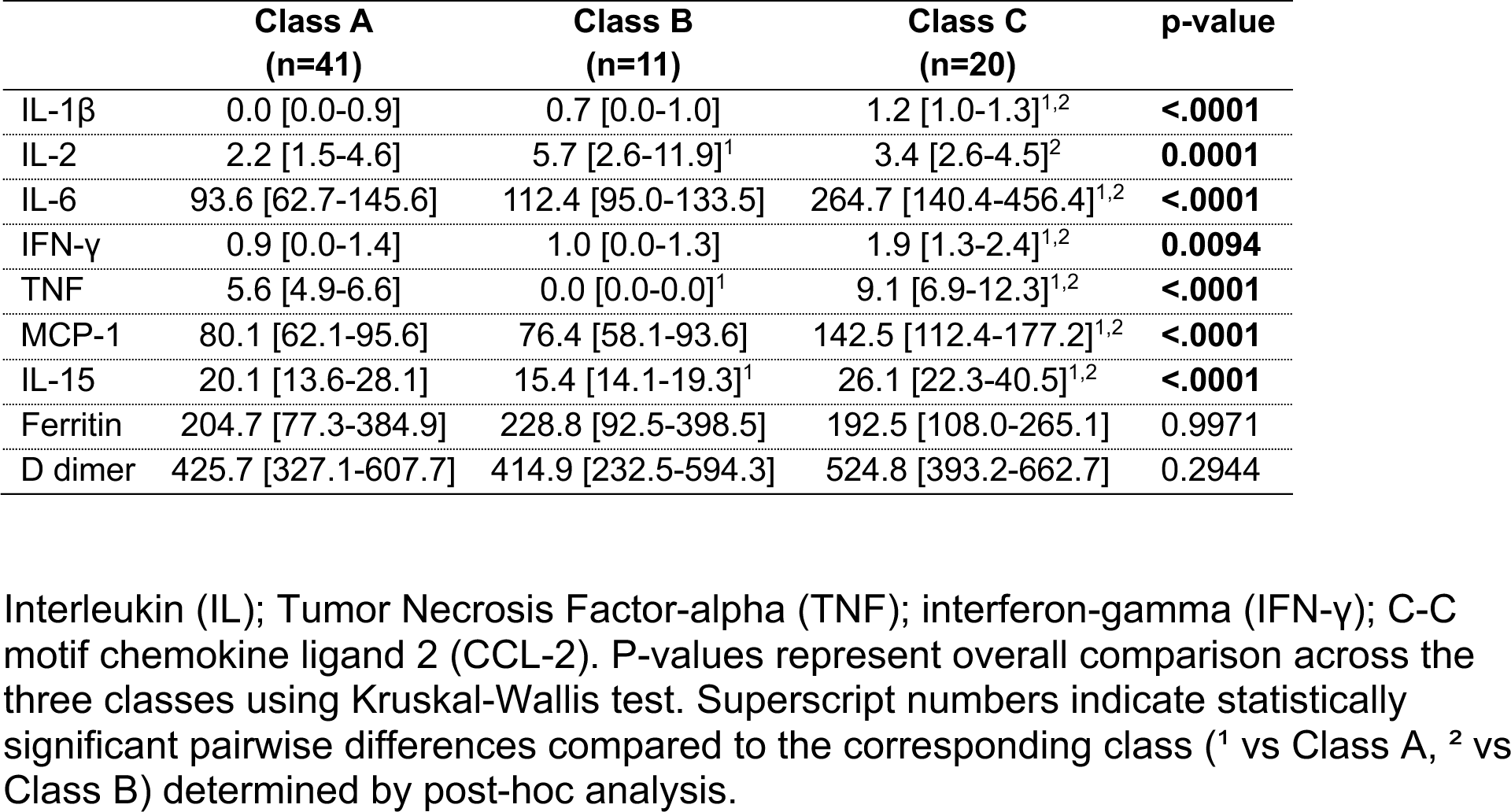
6-hours point-of-care biomarkers concentrations, stratified by classes.

|  | **Class A**<br>**(n=41)** | **Class B**<br>**(n=11)** | **Class C**<br>**(n=20)** | **p-value** |
| --- | --- | --- | --- | --- |
| IL-1β | 0.0 [0.0-0.9] | 0.7 [0.0-1.0] | 1.2 [1.0-1.3]<sup>1,2</sup> | **<.0001** |
| IL-2 | 2.2 [1.5-4.6] | 5.7 [2.6-11.9]<sup>1</sup> | 3.4 [2.6-4.5]<sup>2</sup> | **0.0001** |
| IL-6 | 93.6 [62.7-145.6] | 112.4 [95.0-133.5] | 264.7 [140.4-456.4]<sup>1,2</sup> | **<.0001** |
| IFN-γ | 0.9 [0.0-1.4] | 1.0 [0.0-1.3] | 1.9 [1.3-2.4]<sup>1,2</sup> | **0.0094** |
| TNF | 5.6 [4.9-6.6] | 0.0 [0.0-0.0]<sup>1</sup> | 9.1 [6.9-12.3]<sup>1,2</sup> | **<.0001** |
| MCP-1 | 80.1 [62.1-95.6] | 76.4 [58.1-93.6] | 142.5 [112.4-177.2]<sup>1,2</sup> | **<.0001** |
| IL-15 | 20.1 [13.6-28.1] | 15.4 [14.1-19.3]<sup>1</sup> | 26.1 [22.3-40.5]<sup>1,2</sup> | **<.0001** |
| Ferritin | 204.7 [77.3-384.9] | 228.8 [92.5-398.5] | 192.5 [108.0-265.1] | 0.9971 |
| D dimer | 425.7 [327.1-607.7] | 414.9 [232.5-594.3] | 524.8 [393.2-662.7] | 0.2944 |

Interleukin (IL); Tumor Necrosis Factor-alpha (TNF); interferon-gamma (IFN-γ ); C-C motif chemokine ligand 2 (CCL-2). P-values represent overall comparison across the three classes using Kruskal-Wallis test. Superscript numbers indicate statistically significant pairwise differences compared to the corresponding class (<sup>1</sup> vs Class A, <sup>2</sup> vs Class B) determined by post-hoc analysis.

**Table S2.** Patients’ characteristics stratified by latent biological classes based on 6-hour point-of-care biomarkers.

|  |  | **Class 1**<br>**(n=41)** | **Class 2**<br>**(n=11)** | **Class 3**<br>**(n=20)** | **p** |
| --- | --- | --- | --- | --- | --- |
| Enlistment | Female gender | 19 (46.34) | 4 (36.36) | 9 (45.00) | 0.9842 |
|  | Age at LUTX (years) | 60 [49-64] | 50 [38-61] | 57 [48-60]<sup>1</sup> | 0.9404 |
|  | BMI (kg/m<sup>2</sup>) | 22.1 [18.7-25.5] | 25.5 [20.4-27.7] | 24.2 [19.2-27.0] | 0.4164 |
|  | Diagnosis |  |  |  |  |
|  | Interstitial Lung Disease | 26 (63.41) | 8 (72.73) | 8 (40.00) | 0.3022 |
|  | CF/Bronchiectasis | 9 (21.95) | 3 (27.27) | 3 (15.00) | 0.7889 |
|  | COPD | 5 (12.20) | 0 (0.00) | 4 (20.00) | NA |
|  | Other | 1 (2.44) | 0 (0.00) | 4 (20.00) | NA |
|  | Vascular | 0 (0.00) | 0 (0.00) | 1 (5.00) | NA |
|  | Lung Allocation Score | 44.5 [36.9-50.7] | 37.5 [35.5-43.4] | 41.6 [38.9-45.8] | 0.0729 |
| Donor | Age (years) | 55 [33-60] | 55 [23-62] | 49 [45-56] | 0.7185 |
|  | OTO Score** | 3 [2-5] | 3 [2-4] | 4 [2-5] | 0.4816 |
|  | Cold Ischemia Time (min) | 468 [392-714] | 564 [364-631] | 394 [318-556] | 0.3205 |
|  | Warm Ischemia Time (min) | 73.5 [67.5-84] | 75.5 [68-90] | 71.8 [63-83] | 0.9999 |
|  | DCD (y/n) | 7 (17.50) | 2 (18.18) | 6 (30.00) | 0.6131 |
| Surgical | EVLP (y/n) | 8 (19.51) | 3 (27.27) | 4 (20.00) | 0.9646 |
|  | RBC (units) | 5 [3-9] | 4 [2-10] | 4 [2-6]<sup>1</sup> | 0.0298 |
|  | FFP (units) | 4 [2-5] | 4 [0-10] | 4 [2-6] | 0.4935 |
|  | Blood Components (L) | 2.85 [1.55-4.30] | 2.10 [0.50-7.80] | 2.15 [1.28-3.65] | 0.9999 |
|  | Intraoperative ECMO (y/n) | 22 (53.66) | 5 (45.45) | 12 (60.00) | 0.9562 |

Categorical variables are presented as counts and percentages (n, %), while continuous variables are expressed as medians with interquartile ranges. Abbreviations. BMI: body mass index; CF: Cystic Fibrosis; COPD: Chronic Obstructive Pulmonary Disease; DCD, donor after cardiac death; EVLP, Ex-Vivo Lung Perfusion; RBC, red blood cell; FFP, fresh frozen plasma; ECMO, extracorporeal membrane oxygenation; NA, not applicable.
Notes. \*\* Not calculated for graft undergone EVLP.

**Table S3.**
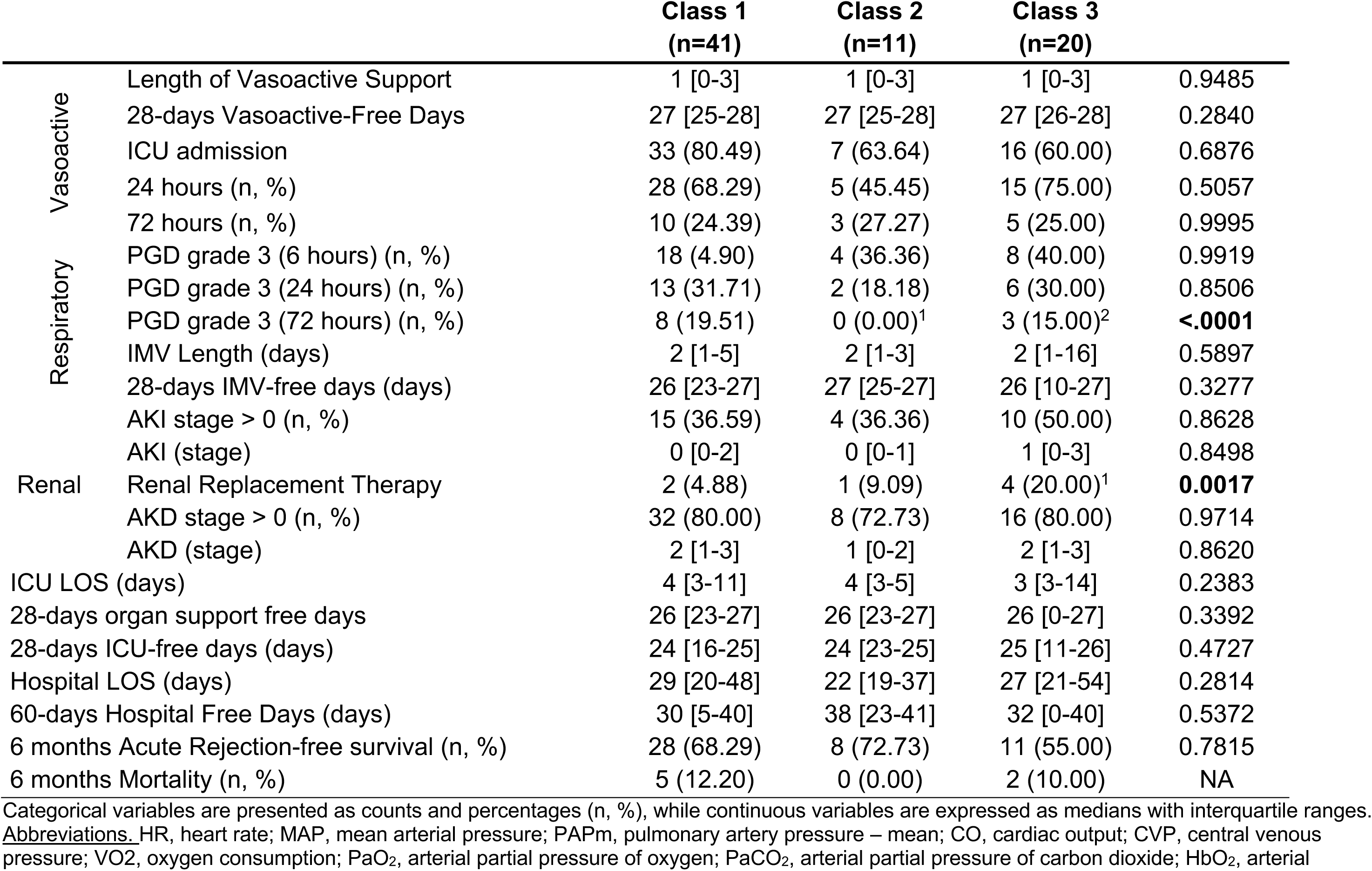
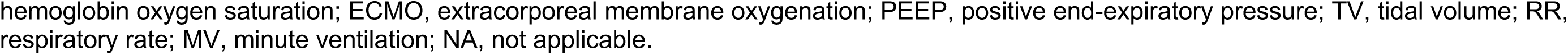
Patients’ outcomes stratified by latent biological classes based on 6-hour point-of-care biomarkers.

|  |  | **Class 1**<br>**(n=41)** | **Class 2**<br>**(n=11)** | **Class 3**<br>**(n=20)** |  |
| --- | --- | --- | --- | --- | --- |
| Vasoactive | Length of Vasoactive Support | 1 [0-3] | 1 [0-3] | 1 [0-3] | 0.9485 |
|  | 28-days Vasoactive-Free Days | 27 [25-28] | 27 [25-28] | 27 [26-28] | 0.2840 |
|  | ICU admission | 33 (80.49) | 7 (63.64) | 16 (60.00) | 0.6876 |
|  | 24 hours (n, %) | 28 (68.29) | 5 (45.45) | 15 (75.00) | 0.5057 |
|  | 72 hours (n, %) | 10 (24.39) | 3 (27.27) | 5 (25.00) | 0.9995 |
| Respiratory | PGD grade 3 (6 hours) (n, %) | 18 (4.90) | 4 (36.36) | 8 (40.00) | 0.9919 |
|  | PGD grade 3 (24 hours) (n, %) | 13 (31.71) | 2 (18.18) | 6 (30.00) | 0.8506 |
|  | PGD grade 3 (72 hours) (n, %) | 8 (19.51) | 0 (0.00)<sup>1</sup> | 3 (15.00)<sup>2</sup> | **<.0001** |
|  | IMV Length (days) | 2 [1-5] | 2 [1-3] | 2 [1-16] | 0.5897 |
|  | 28-days IMV-free days (days) | 26 [23-27] | 27 [25-27] | 26 [10-27] | 0.3277 |
| Renal | AKI stage > 0 (n, %) | 15 (36.59) | 4 (36.36) | 10 (50.00) | 0.8628 |
|  | AKI (stage) | 0 [0-2] | 0 [0-1] | 1 [0-3] | 0.8498 |
|  | Renal Replacement Therapy | 2 (4.88) | 1 (9.09) | 4 (20.00)<sup>1</sup> | **0.0017** |
|  | AKD stage > 0 (n, %) | 32 (80.00) | 8 (72.73) | 16 (80.00) | 0.9714 |
|  | AKD (stage) | 2 [1-3] | 1 [0-2] | 2 [1-3] | 0.8620 |
| ICU LOS (days) |  | 4 [3-11] | 4 [3-5] | 3 [3-14] | 0.2383 |
| 28-days organ support free days |  | 26 [23-27] | 26 [23-27] | 26 [0-27] | 0.3392 |
| 28-days ICU-free days (days) |  | 24 [16-25] | 24 [23-25] | 25 [11-26] | 0.4727 |
| Hospital LOS (days) |  | 29 [20-48] | 22 [19-37] | 27 [21-54] | 0.2814 |
| 60-days Hospital Free Days (days) |  | 30 [5-40] | 38 [23-41] | 32 [0-40] | 0.5372 |
| 6 months Acute Rejection-free survival (n, %) |  | 28 (68.29) | 8 (72.73) | 11 (55.00) | 0.7815 |
| 6 months Mortality (n, %) |  | 5 (12.20) | 0 (0.00) | 2 (10.00) | NA |

Categorical variables are presented as counts and percentages (n, %), while continuous variables are expressed as medians with interquartile ranges. Abbreviations: HR, heart rate; MAP, mean arterial pressure; PAPm, pulmonary artery pressure – mean; CO, cardiac output; CVP, central venous pressure; VO<sub>2</sub>, oxygen consumption; PaO<sub>2</sub>, arterial partial pressure of oxygen; PaCO<sub>2</sub>, arterial partial pressure of carbon dioxide; HbO<sub>2</sub>, arterial
hemoglobin oxygen saturation; ECMO, extracorporeal membrane oxygenation; PEEP, positive end-expiratory pressure; TV, tidal volume; RR, respiratory rate; MV, minute ventilation; NA, not applicable.

**Figure S2.**
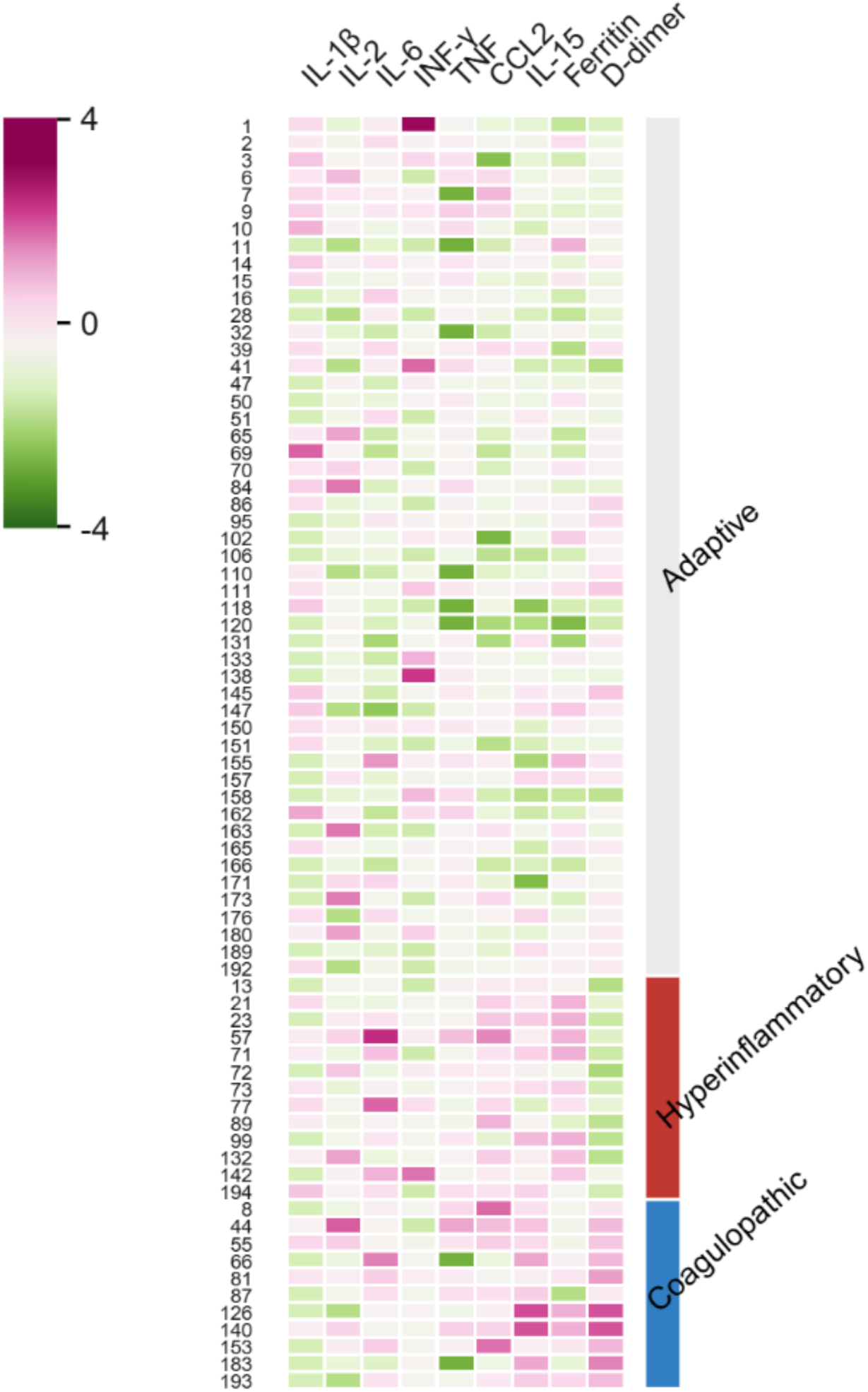
Individual plasma concentrations of biomarkers used for detecting phenotypes in the latent profile analysis. Heatmap of individual biomarker concentrations at 36 hours stratified by phenotypes. Columns represent biomarkers used for the latent profile analysis; rows represent individual subjects. Color scale indicates standardized concentrations: pink represents higher concentrations and green represents lower concentrations relative to the cohort mean.

**Table S4.**
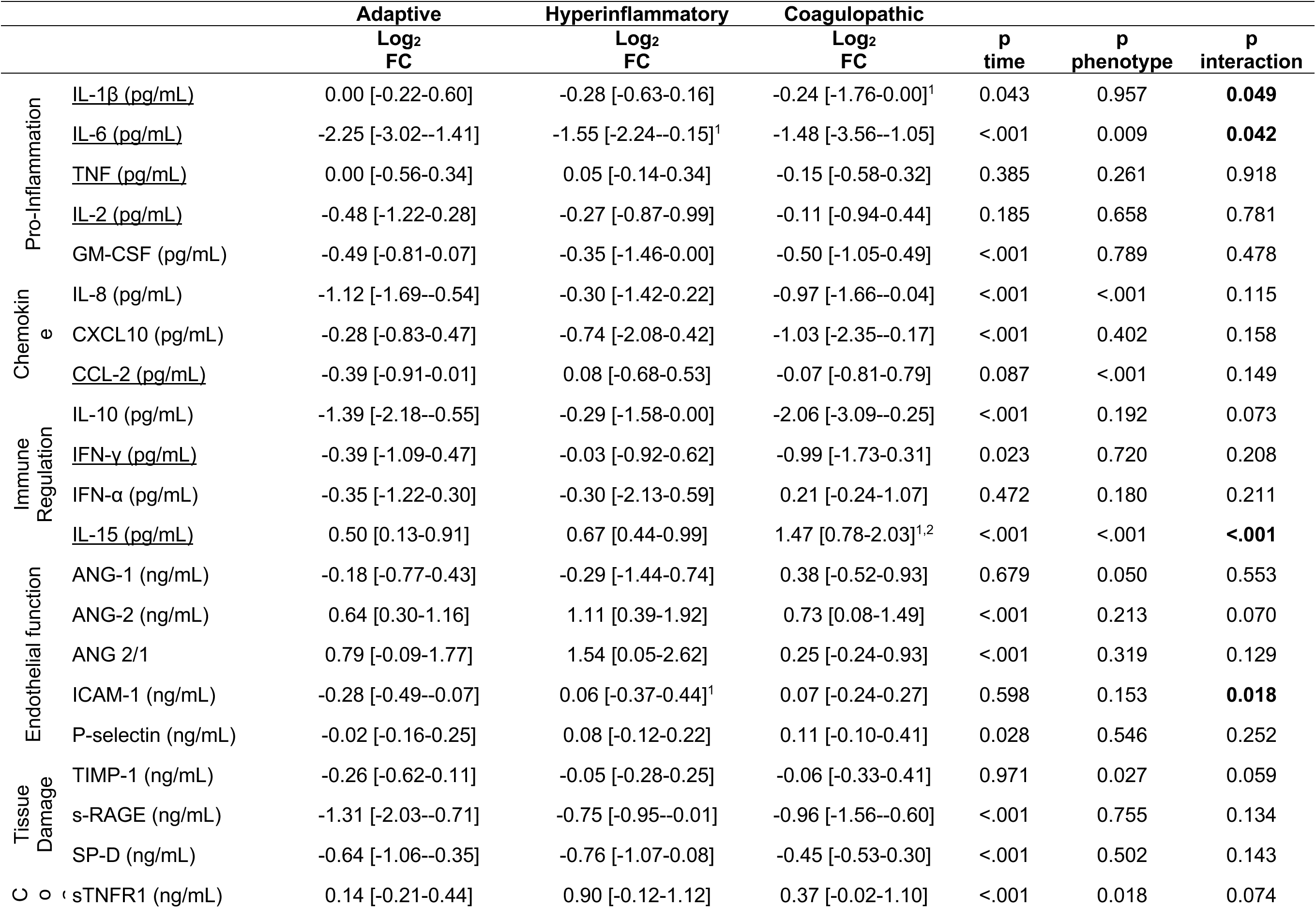
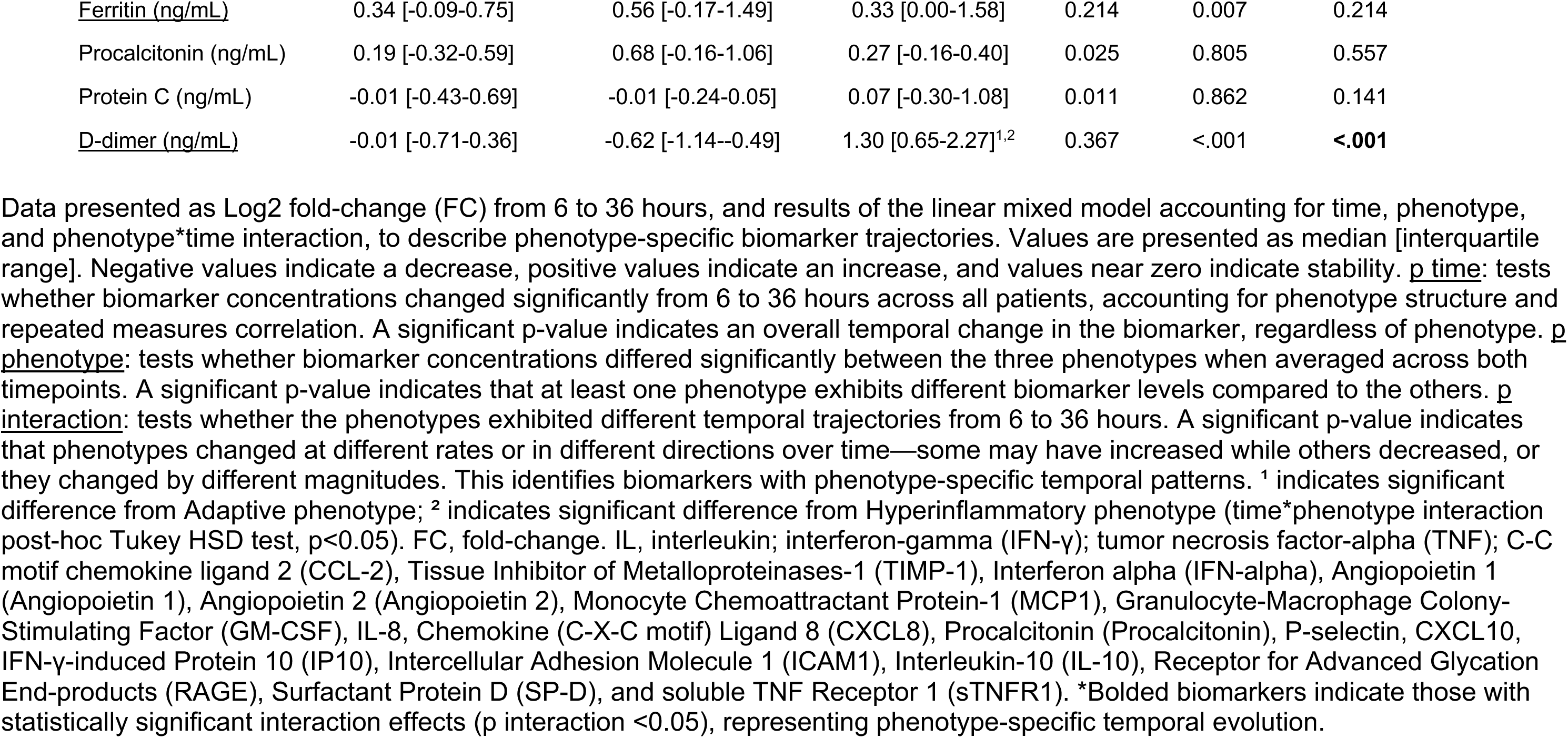
Longitudinal biomarker changes (from 6 to 36 hours) stratified by phenotype.

## Bibliography

1. Weill D, Benden C, Corris PA, et al.: A consensus document for the selection of lung transplant candidates: 2014--an update from the Pulmonary Transplantation Council of the International Society for Heart and Lung Transplantation. J Heart Lung Transplant 34: 1–15, 2015

2. Snell GI, Yusen RD, Weill D, et al.: Report of the ISHLT Working Group on Primary Lung Graft Dysfunction, part I: Definition and grading—A 2016 Consensus Group statement of the International Society for Heart and Lung Transplantation J Hear Lung Transplant 36: 1097–1103, 2017

3. Takahashi T, Terada Y, Pasque MK, et al.: Outcomes of Extracorporeal Membrane Oxygenation for Primary Graft Dysfunction After Lung Transplantation. Ann Thorac Surg 115: 1273–1280, 2023

4. Shah RJ, Diamond JM, Cantu E, et al.: Latent class analysis identifies distinct phenotypes of primary graft dysfunction after lung transplantation Chest 144: 616–622, 2013

5. Scaravilli V, Guzzardella A, Madotto F, et al.: Hemodynamic failure and graft dysfunction after lung transplant: A possible clinical continuum with immediate and long-term consequences. Clin Transplant 37: e15122, 2023

6. Scaravilli V, Merrino A, Bichi F, et al.: Longitudinal assessment of renal function after lung transplantation for cystic fibrosis: transition from post-operative acute kidney injury to acute kidney disease and chronic kidney failure J Nephrol 35: 1885–1893, 2022

7. Bos LD, Schouten LR, Van Vught LA, et al.: Identification and validation of distinct biological phenotypes in patients with acute respiratory distress syndrome by cluster analysis Thorax 72: 876–883, 2017

8. Sinha P, Delucchi KL, Thompson BT, et al.: Latent class analysis of ARDS subphenotypes: a secondary analysis of the statins for acutely injured lungs from sepsis (SAILS) study. Intensive Care Med 44: 1859–1869, 2018

9. Seymour CW, Kennedy JN, Wang S, et al.: Derivation, Validation, and Potential Treatment Implications of Novel Clinical Phenotypes for Sepsis. JAMA 321: 2003–2017, 2019

10. Scaravilli V, Turconi G, Colombo SM, et al.: Early serum biomarkers to characterise different phenotypes of primary graft dysfunction after lung transplantation: a systematic scoping review. ERJ open Res 10, 2024

11. Loupy A, Sablik M, Khush K, Reese PP: Advancing patient monitoring, diagnostics, and treatment strategies for transplant precision medicine Lancet 406: 389–402, 2025

12. Vandenbroucke JP, von Elm E, Altman DG, et al.: Strengthening the Reporting of Observational Studies in Epidemiology (STROBE): explanation and elaboration. PLoS Med 4: e297, 2007

13. Fumagalli J, Rosso L, Gori F, et al.: Early pulmonary function and mid-term outcome in lung transplantation after ex-vivo lung perfusion - a single-center, retrospective, observational, cohort study. Transpl Int 33: 773–785, 2020

14. Valenza F, Rosso L, Gatti S, et al.: Extracorporeal lung perfusion and ventilation to improve donor lung function and increase the number of organs available for transplantation. Transplant Proc 44: 1826–9, 2012

15. Scaravilli V, Morlacchi LC, Merrino A, et al.: Intraoperative extracorporeal membrane oxygenation for lung transplantation in cystic fibrosis patients: Predictors and impact on outcome. J Cyst Fibros 19: 659–665, 2020

16. Scaravilli V, Scansani S, Grasso A, et al.: Right Ventricle Dysfunction in Patients With Adult Cystic Fibrosis Enlisted for Lung Transplant. Transplant Proc 53: 260–264, 2021

17. Scaravilli V, Merrino A, Bichi F, et al.: Longitudinal assessment of renal function after lung transplantation for cystic fibrosis: transition from post-operative acute kidney injury to acute kidney disease and chronic kidney failure J Nephrol, 2022

18. Scaravilli V, Scansani S, Meani P, et al.: Right ventricle free wall longitudinal strain screening of lung transplant candidates. PLoS One 19: e0314235, 2024

19. Kanou T, Nakahira K, Choi AM, et al.: Cell-free DNA in human ex vivo lung perfusate as a potential biomarker to predict the risk of primary graft dysfunction in lung transplantation J Thorac Cardiovasc Surg 162: 490-+, 2021

20. Oto T, Levvey BJ, Whitford H, et al.: Feasibility and Utility of a Lung Donor Score: Correlation With Early Post-Transplant Outcomes Ann Thorac Surg 83: 257–263, 2007

21. Fumagalli J, Rosso L, Gori F, et al.: Early pulmonary function and mid-term outcome in lung transplantation after ex-vivo lung perfusion – a single-center, retrospective, observational, cohort study Transpl Int 33: 773–785, 2020

22. KDIGO Clinical Practice Guideline for Acute Kidney Injury. 2012 Mar;2(1):8–12.: Kidney Int Suppl Mar: 8–12, 2012

23. Chawla LS, Bellomo R, Bihorac A, et al.: Acute kidney disease and renal recovery: Consensus report of the Acute Disease Quality Initiative (ADQI) 16 Workgroup Nat Rev Nephrol 13: 241–257, 2017

24. Renaud-Picard B, Koutsokera A, Cabanero M, Martinu T: Acute Rejection in the Modern Lung Transplant Era. Semin Respir Crit Care Med 42: 411–427, 2021

25. Ong T, McClintock DE, Kallet RH, Ware LB, Matthay MA, Liu KD: Ratio of angiopoietin-2 to angiopoietin-1 as a predictor of mortality in acute lung injury patients. Crit Care Med 38: 1845–51, 2010

26. Lissa CJ Van, Anadria D: Recommended Practices in Latent Class Analysis Using the Open-Source R-Package tidySEM Recommended Practices in Latent Class Analysis Using the Open-Source Struct Equ Model A Multidiscip J 31: 526–534, 2024

27. Bolck A, Croon M, Hagenaars J: Estimating Latent Structure Models with Categorical Variables: One-Step Versus Three-Step Estimators Polit Anal 12: 3–27, 2004

28. Scaravilli V, Guzzardella A, Madotto F, et al.: Hemodynamic failure and graft dysfunction after lung transplant: A possible clinical continuum with immediate and long-term consequences Clin Transplant: 1–10, 2023

29. Shah RJ, Bellamy SL, Localio AR, et al.: A panel of lung injury biomarkers enhances the definition of primary graft dysfunction (PGD) after lung transplantation J Hear Lung Transplant 31: 942–949, 2012

30. Pottecher J, Roche AC, Degot T, et al.: Increased Extravascular Lung Water and Plasma Biomarkers of Acute Lung Injury Precede Oxygenation Impairment in Primary Graft Dysfunction After Lung Transplantation Transplantation 101: 112–121, 2017

31. Chacon-Alberty L, Kanchi RS, Ye SB, et al.: Plasma protein biomarkers for primary graft dysfunction after lung transplantation: a single-center cohort analysis Sci Rep 12, 2022

32. Yang W, Cerier EJ, Núñez-Santana FL, et al.: IL-1β–dependent extravasation of preexisting lung-restricted autoantibodies during lung transplantation activates complement and mediates primary graft dysfunction J Clin Invest 132

33. Moreno I, Mir A, Vicente R, et al.: Analysis of Interleukin-6 and Interleukin-8 in Lung Transplantation: Correlation With Nitric Oxide Administration Transplant Proc 40: 3082–3084, 2008

34. Verleden SE, Martens A, Vandermeulen E, et al.: The association of IL-6 and IL-8 within 72 hours post-transplant and short and long term outcome, 2017.

35. Allen JG, Lee MT, Weiss ES, Arnaoutakis GJ, Shah AS, Detrick B: Preoperative Recipient Cytokine Levels Are Associated With Early Lung Allograft Dysfunction Ann Thorac Surg 93: 1843–1849, 2012

36. Hoffman SA, Wang L, Shah C V, et al.: Plasma Cytokines and Chemokines in Primary Graft Dysfunction Post-Lung Transplantation Am J Transplant 9: 389–396, 2009

37. Chacon-Alberty L, Ye S, Elsenousi A, et al.: Effect of intraoperative support mode on circulating inflammatory biomarkers after lung transplantation surgery Artif Organs 47: 749–760

38. Bharat A, Kuo E, Steward N, et al.: Immunological link between primary graft dysfunction and chronic lung allograft rejection Ann Thorac Surg 86: 189–197, 2008

39. Du J, Yang H, Zhang D, et al.: Structural basis for the blockage of IL-2 signaling by therapeutic antibody basiliximab. J Immunol 184: 1361–8, 2010

40. Calfee CS, Delucchi KL, Sinha P, et al.: Acute respiratory distress syndrome subphenotypes and differential response to simvastatin: secondary analysis of a randomised controlled trial Lancet Respir Med 6: 691–698, 2018

41. Hong SK, Han D, Lee SK, et al.: Short-term therapy with anti-ICAM-1 monoclonal antibody induced long-term liver allograft survival in nonhuman primates Am J Transplant 21: 2978–2991, 2021

